# The genomic landscape of syndromic and non-syndromic hearing loss within the 100,000 Genomes Project cohort

**DOI:** 10.1101/2025.02.06.25321804

**Authors:** Letizia Vestito, Damian Smedley, Valentina Cipriani, Gudrun E Moore, Philip Stanier, Michael R Bowl, Sally J Dawson, Emma Clement, Maria Bitner-Glindzicz

## Abstract

**Objective:** This study aims to describe the genetic landscape of syndromic and non-syndromic hearing loss (HL) in the UK population using data from the 100,000 Genomes Project (100kGP).

**Design:** Cohort study

**Setting:** NHS England

**Participants:** 2,271 families with syndromic and non-syndromic HL recruited to the 100kGP rare disease programme between 2013 and 2018. Participants with at least one Human Phenotype Ontology (HPO) term descendant of the term “Hearing impairment” (HP:0000365) were included; this equated to 5,488 individuals, comprising 2,762 affected individuals and 2,726 unaffected relatives.

**Main outcome measure:** Diagnostic rate and prevalence of different gene diagnoses by auditory phenotype identified by whole genome sequencing.

**Results:** The overall diagnostic yield was conservatively estimated at 27.5% (625/2271), with diagnoses identified in 273 different genes. Common causative genes included *USH2A, GJB2, COL1A1* and *MYO15A*, accounting for approximately 20% of the diagnoses. This diagnostic rate excludes variants of uncertain significance (VUS), variants in genes where HL cannot be confidently attributed to the identified variant, or those still awaiting confirmation. The inclusion of these categories would increase the diagnostic yield to 39.6%. This work describes the 100kGP standard pipeline and supplementary analyses that include the use of Exomiser. Stratification of the cohort allowed quantification of the likelihood of genetic diagnosis with specific phenotypic combinations and identification of positive predictors for a genetic diagnosis by auditory phenotype. A statistically significant increase in diagnostic rate was reported for those with congenital (33.2%), bilateral (27%), and high-frequency (32.4%) hearing subtypes. Furthermore, in patients with HPO terms restricted to the auditory system alone, around 40% of diagnoses were attributed to genes that might have a broader syndromic phenotype (non-syndromic mimics). A high diagnostic yield (56%) was seen in patients with ear and eye abnormalities, largely driven by genes associated with Usher and Wolfram syndrome.

**Conclusion:** In conclusion, this study offers valuable insights into the complex genomic and phenotypic architecture of both syndromic and non-syndromic HL, which has the potential to improve diagnostic pipelines and inform clinical care.

## Introduction

Hearing loss (HL) is a highly heterogeneous sensory impairment with a prevalence of around 1/500 newborn babies [1]. HL may be broadly classified according to underlying aetiology and the specific part of the auditory system affected [2]. Pathology involving the external and middle ear, which convey the sound and transmit the sound wave to the inner ear typically causes conductive HL. Inner ear pathology interfering with the transformation of the mechanical sound stimulus into a neural impulse via the auditory nerve pathways causes sensorineural HL. A combination of both types is called mixed HL. HL can be further defined based on the frequencies involved, severity, progression or age of onset [3, 4] It has been estimated that 80% of prelingual HL is likely to have a genetic cause, while in the remaining 20%, the HL is likely to be acquired or have an environmental aetiology [5]. Approximately 80% of genetic HL cases are classified as non-syndromic, with no additional features outside the auditory system. The remaining 20% are considered syndromic [6]. The Hereditary Hearing Loss database reports 124 non-syndromic genes [7] and over 400 syndromes involving HL are described in OMIM [8, 9]. This reflects the complexity of the genomic and phenotypic architecture of HL. The reported diagnostic yield of genetic testing in patients with HL is highly variable, between 20% and 50% [10-20]. Several factors may contribute to this, including ancestry, marked variances in inclusion/exclusion criteria used across different projects, pre-screening for known prevalent genes like *GJB2*, and the methodology used. However, the respective contributions of less prevalent genes to the frequency and type of genetic deafness remain largely unknown. A greater understanding of these contributions is necessary to translate genetic technology advances into improvements in diagnosis and future personalised therapeutic strategies.

Historically, the diagnosis of HL has relied on targeted panel-based genetic tests, which include a limited set of known genes associated with HL. More recently, next-generation sequencing technologies enhanced the transition from panel-based tests to whole exome (WES) and whole genome sequencing (WGS) in the diagnostic setting. The 100,000 Genomes Project (100kGP) [21, 22] undertook WGS for National Health Service (NHS) patients with cancer or a wide range of rare diseases, including hearing phenotypes, enrolled through one of the NHS Genomic Medicine Centres (GMCs) across the UK [21, 22]. This allowed a unique opportunity to catalogue rare genomic sequence variants and correlate them with their patient’s hearing phenotypes.

This work’s primary objective is to describe the genetic landscape of syndromic and non-syndromic HL in the UK population by analysing the 100kGP data. Here, we report the findings from 2,271 families with HL recruited into the 100kGP rare disease programme, one of the largest cohorts examined to date.

## Methods

### Patient cohort and Ethical Approval

The 100,000 Genomes Project (100kGP) is a UK Government project launched in 2013 and coordinated by Genomics England, a wholly owned company of the Department of Health and Social Care. The project aims to apply whole-genome sequencing to the study of rare diseases, cancers, and infections in a national healthcare setting. After ethical approval (National Research Ethics approvals 14/EE/1112 and 13/EE/032), NHS rare disease patients (probands and family members) were recruited to the 100kGP Rare Disease programme (pilot or main) through one of 13 Genomic Medicine Services (GMCs) around the UK. All participants provided written informed consent. Probands were recruited according to disease-specific clinical eligibility criteria established by Genomics England; information about recruitment guidelines and exclusion/ inclusion criteria can be found at www.genomicseducation.hee.nhs.uk/eligibility-wheels.

Patient-specific phenotypic information exists as two main sets of data: the ‘disease category’ under which patients were recruited and the Human Phenotype Ontology (HPO) terms [23, 24] selected by the clinician at the time of recruitment. 2271 probands with at least one of the 37 HPO term descendants of “Hearing impairment” (HP:0000365) were included in the analysis. The full list of HPO terms is available in <u>Supplementary Table 1</u>. Data from family members was included in the study where available. Of these probands, 34.1% (774/2271) were recruited under the “hearing and ear disorders” disease group, for whom HL likely manifests as the predominant clinical phenotype, which constitutes the “*HED”* cohort. Seven specific HL disease subcategories were defined as follows: “congenital hearing impairment”, “autosomal dominant deafness”, “bilateral microtia”, “ear malformations with hearing impairment”, “auditory neuropathy spectrum disorder”, “familial hemifacial microsomia” and “familial Meniere disease”. Prior genetic testing for common genetic causes (*GJB2, GJB6*, or *SLC26A4)* was a recommended prerequisite to recruitment into the HL disease categories. Further information on recruiting disease categories and subcategories can be found in <u>Table 1: Overview of cohorts</u>’ <u>demographics</u>.

### Patient and Public Involvement

The Participant Panel is a voluntary advisory group established to represent the interests of thousands of individuals whose data is held by Genomics England in the National Genomics Research Library (NGRL), which includes the 100kGP. The panel collaborate with Genomics England to ensure the diverse voices of participants, patients, and their families are heard and understood. They influence decisions about project design and the use of participants’ sequenced genomes and associated health data. Project RR140 was registered as part of the Hearing and Sight Genomics England Clinical Interpretation Partnership (GeCIP), reviewed and accepted by Genomics England through an Access Review Committee and the Ethics Advisory Committee.

### WGS and sequencing pipeline

Samples, either as DNA extracted from whole blood or as whole blood EDTA samples, were processed at the National Institutes for Health Research BioResource Laboratory in Cambridge. They underwent testing to ensure adequate DNA concentration and were quality-controlled for degradation and purity. WGS was conducted using the Illumina TruSeq DNA PCR-free sample preparation kit from Illumina on the HiSeqX sequencer, achieving an average depth of 32× across the human genome. Sequencing reads were aligned to the GRCh37 or GRCh38 human genome reference using Isaac Genome Alignment Software. Additional information can be found at [21, 22]. Family-based variant calling for single-nucleotide variants (SNVs) and small insertions or deletions (indels) was performed using the Platypus variant caller [25]. Copy Number Variants (CNVs) and Structural Variants (SVs) were called using Canvas [26] and Manta [27]. Manta was employed to detect CNVs and SVs using read-pair and split-read detection, while Canvas was utilised to identify CNVs larger than 10 kb by detecting gaps in read depth.

### 100kGP interpretation pipeline

The standard 100kGP analysis pipeline was disease-specific, panel-based and driven by clinical indication. Panel content and gene classification criteria are described in PanelApp (https://panelapp.genomicsengland.co.uk) [28]. This pipeline systematically evaluated variants within genes demonstrating robust evidence of association with specific diseases (designated as green genes), indicating their potential diagnostic relevance. Conversely, genes exhibiting conflicting evidence (amber genes) or insufficient evidence for pathogenicity (red genes) were initially excluded from the analysis. The analysis pipeline first filters out variants that do not follow the expected inheritance pattern within the family, do not impact protein-coding sequences, or variants with high frequencies in external and local datasets. These filtered variants are categorised as “Untiered” or “Tier null.” Filtered variants in green PanelApp genes typically fall into Tier 1 or Tier 2. Tier 1 candidates include loss-of-function or *de novo* variants affecting green genes within the applied panels, while Tier 2 comprises other variant types, such as missense variants affecting these genes. In the analysis of the 100kGP data, variants in Tier 1 and Tier 2 were assessed by clinical scientists in each GMC according to the guidelines of the Association for Clinical Genomic Science (ACGS) [21, 22]. Tier 3 variants were not routinely analysed through the standard Genomics England pipeline and include rare protein-altering variants that segregate within families and are found in genes categorised as ‘Amber’ or ‘Red’ in the panel, as well as in genes outside the applied panel. Variants deemed pathogenic or likely pathogenic were reported and fed back to families via recruiting clinicians. Each proband’s diagnostic status was categorised as follows: ‘diagnosed’ status indicates that the proband’s clinical phenotype is fully accounted for by an identified pathogenic or likely pathogenic variant; ‘partially diagnosed’ denotes that the identified variant only partially explains the proband’s clinical features, suggesting the involvement of additional genetic or environmental factors, ‘potential candidate awaiting confirmation’ applies when a pathogenic or likely pathogenic variant has been identified but requires validating in a diagnostic lab, ‘VUS’ indicates that the proband has a variant of uncertain significance in a candidate gene, though its clinical significance remains unresolved, ‘undiagnosed’ applies to probands for whom no diagnostic or candidate variants have been identified, leaving their condition without a genetic explanation. Each of these categories was then further divided based on whether the candidate gene is a green gene in the HL panel (‘HL explained’) or not.

### Exomiser and additional analysis

Exomiser software v12.1.0 [29, 30] was run on the 2271 rare disease families recruited to the 100kGP having at least one HPO term descendant of the “Hearing impairment” HPO term (HP:0000365) using standard Exomiser settings, which include different variant frequency threshold per mode of inheritance (MAF <0.1% or 2% for compound-heterozygotes or mitochondrial) applied to all of the following reference databases: 100,000 Genomes Project reference samples, 1000 Genomes, ESP, TOPMed, UK10K, ExAC and gnomAD. The in silico pathogenicity predictors used were REVEL and MVP, which encompass many other pathogenicity predictors, e.g. Polyphen2, SIFT and MutationTaster from dbNSFP. The full list of HPO terms is available in <u>Supplementary Table 1</u>. The dataset consists of a total of 2,102 probands who are part of the main program v18, and an additional 169 probands from the pilot v3. Exomiser scores and ranks the rare, segregating, predicted pathogenic variants that pass the filters based on the predicted pathogenicity score(s), MAF and similarity of the patient phenotypes to reference genotype to phenotype knowledge from known disease-gene associations, model organisms and protein-protein networks. In a proportion of cases, CNV/SV analyses were performed using Exomiser or SVrare [31]. The coverage quality for the SNV or CNV/SV was assessed by inspecting the genomic region using Integrative Genome Viewer (IGV) software [32] from the Binary Alignment Map (BAM) file available per each proband within the Genomics England Research Environment. Findings of diagnostic relevance were validated using SNParray by recruiting GMCs. The cohorts’ demographics, presented in Table 1, were obtained via Labkey in the Genomics England Research Environment. Genetic ancestry inference was performed by Genomics England via principal component analysis. A random forest model was subsequently trained to predict ancestry across five super-populations (i.e. European, African, Admixed American, South Asian, East Asian), with individuals assigned to ancestries based on a probability threshold of > 0.8 (https://re-docs.genomicsengland.co.uk/ancestry_inference/).

### Data stratification and visualisation

The hierarchical structure of HPO terms facilitated computational distinction between syndromic and non-syndromic cases. Non-syndromic patients were defined as those who exhibited HPO terms solely related to the “Abnormality of the ear” branch of the hierarchical tree. On the other hand, syndromic patients were classified as individuals with additional HPO terms from different branches, such as “Abnormality of the eye” or “Abnormality of the nervous system”. This enabled the computational stratification of the cohort into syndromic and non-syndromic groups.

The use of HPO terms also allowed further cohort stratification based on the classification of their HL, including the type, severity, onset, progression, laterality, and frequency. Gene ontology enrichment analysis and visualisation were performed using GOnet [33]. R/v3.6.1 [34] was used to generate tables and figures. A two-sided Fisher’s exact test was used to compare diagnostic yields in the stratified cohort.

## Results

### Description of the HL patient cohort

The cohort of patients described in this work includes 2271 rare disease families recruited to the 100kGP, with the proband having at least one HPO term descendant of the term “Hearing impairment” (HP:0000365). This group comprises 5488 individuals, with 2762 affected individuals and 2726 unaffected relatives. As indicated in <u>Table 1: Overview of cohorts’ demographics</u>, 48% of the probands (1090 families) were recruited as trios (proband and both biological parents).). Additional 565 probands were recruited as singletons (24.9%) and 448 as duos (proband and one family member, typically one parent) (19.7%). A smaller number were recruited as quadruplets (146 probands, 6.4%) or with five or more family members (22 probands, 1%). The age range of the probands was 0 to 94 years old, with a median age of 20 years and a mean age of 29 years. The self-reported ethnic categories were available for 76% of the cohort. Of the total cohort, 53.5% identified as White British (70.4% within the subset with available data), and 5.6% identified as belonging to another White background (7.4% within the subset with available data). The next most common ethnic groups were Pakistani, making up 5% of the overall cohort (6.6% within the subset with available data), and Indian, representing 2.7% of the overall cohort (3.5% within the subset with available data). The cohort was also classified into six inferred ancestry groups (see Methods). The majority of participants were of European ancestry (n = 1465, 64.5%), followed by South Asian ancestry (n = 257, 11.3%). A smaller proportion of participants were classified as African (n = 30, 1.3%), East Asian (n = 13, 0.6%), and Admixed American (n = 8, 0.4%), the remaining were unassigned in terms of ancestry (n = 498, 21.9%). The inferred ancestry proportions in this cohort are largely similar to those observed in the broader Rare Disease Programme, with a few differences. Specifically, there is a 4.8% reduction in the proportion of individuals with European ancestry and a 3.1% increase in the representation of individuals with South Asian ancestry. Smaller variations are observed in the African and East Asian groups, with respective changes of -1.1% and +0.1%; while the proportion of individuals with American ancestry remains unchanged. These shifts suggest that this cohort exhibits slightly greater ancestral diversity in comparison to the broader programme. Consanguinity was reported in 7.4% of the cohort, with a further 0.8% identified as possibly consanguineous. This represents a 2.4% increase in the total proportion of consanguinity compared to the broader Rare Disease Programme. The higher proportion of confirmed consanguinity in this cohort could suggest a greater prevalence of genetic conditions with autosomal recessive inheritance patterns, which are more likely to manifest in populations with higher rates of consanguineous relationships or may also reflect differences in the ancestry composition of the cohort, where consanguinity tends to be more common in certain population groups.

The largest proportion of patients (34.1%) in this study were recruited under the disease group “hearing and ear disorders” (n = 774 probands). Other common disease groups contributing to the cohort were “neurology and neurodevelopmental disorders” with 25.5% (578 probands), “ophthalmological disorders” with 6.3% (142 probands), “ultra-rare disorders” with 5.9% (135 probands), and “metabolic disorders” with 5.1% (115 probands). The HL cohort included patients recruited under various disease categories as long as they exhibited HPO terms describing hearing impairment, therefore, a higher proportion of syndromic cases was expected. Based on the HPO terms classification method, 78% (1771 individuals) of the HL cohort were considered syndromic, while the remaining 22% (500 individuals) were classified as non-syndromic. A detailed description of the cohort is available in <u>Table 1: Overview of cohorts</u>’ <u>demographics</u>.

The HPO terms were also used to assess the characteristics of HL, such as its type, severity, onset, progression, laterality, and frequency. Using the HPO terms was instrumental in measuring and comparing the diagnostic yield within specific sub-categories of HL. The most predominant auditory phenotypes identified in the cohort were sensorineural HL, bilateral presentation, and congenital. Information about the type of HL was available for 70.2% of the cohort, with sensorineural HL observed in 57.8% of those cases. Onset information was reported for 29.8% of the cohort, of which 16.3% had congenital onset. Laterality data was available for 38% of the cohort, with 32.5% exhibiting bilateral HL.

A subgroup analysis was also undertaken, focusing on 774 families recruited under the “hearing and ear disorders” disease group. This group comprises 2039 individuals, with 1067 affected individuals and 972 unaffected relatives (*HED cohort)*. The cohorts’ demographics are available in <u>Table 1: Overview of cohorts’ demographics</u>.

### Overall diagnostic yield and genomic architecture

The overall diagnostic yield within the HL cohort was 27.5% (625/2271). This yield only considered patients whose genetic diagnosis fully explains the clinical presentation (579 probands) and partially diagnosed individuals whose HL is attributed to a diagnosis in a gene marked as green in the HL PanelApp panels (46 probands) [28]. This highly conservative diagnostic yield estimate excludes an additional 66 probands with partial diagnoses where the implicated gene is not a green gene in the HL PanelApp panels. It is noteworthy that mutations in some of the genes within this excluded subset have been previously reported in patients with HL [7, 9, 35] but with insufficient evidence to allow us to confidently attribute the HL phenotype to the variant identified. There are an additional 39 probands with candidate variants awaiting confirmation and 169 with VUS in green HL genes on PanelApp, for which we could not ascertain the causality of the hearing impairment from the information available. The inclusion of these categories would increase the diagnostic yield to 39.6%. The summary statistics of the 625 diagnostic findings within the HL cohort can be found in Figure 1A and <u>Table 2: Diagnostic overview</u>.

**Figure 1:**
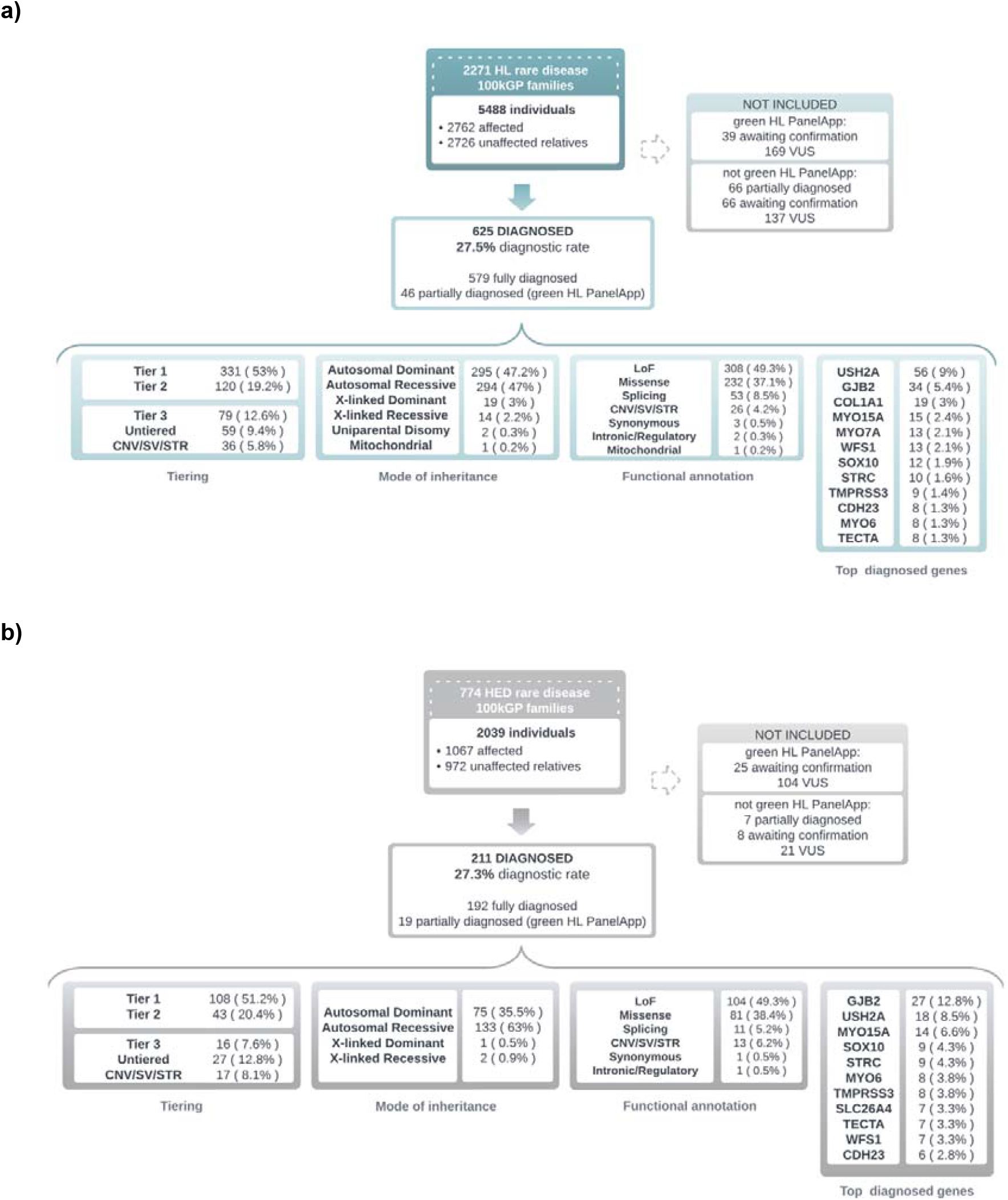
**a)** Summary statistics of the HL cohort. The cohort includes 2271 rare disease families with at least one of the 37 HPO term descendants of “Hearing impairment” (HP:0000365). 625 have been diagnosed and therefore contribute to the diagnostic yield of 27.5%. **b)** Summary statistics of the HED cohort. The cohort includes 774 rare disease families recruited under the disease group “hearing and ear disorders”. 211 have been diagnosed and therefore contribute to the diagnostic yield of 27.3%. Additional stats, including tiering, mode of inheritance, functional annotation and top-diagnosed genes, are available for both cohorts.

72.2% of the overall diagnoses were identified in Tier 1 or Tier 2 variants and were screened through the standard Genomics England pipeline. The remaining 27.8% (equating to 174 diagnoses) includes rare protein-altering variants that segregated within families and were found in genes categorised as ‘Amber’ or ‘Red’ in the applied panels, or in genes not in the applied panels (Tier 3 variants), which constitute the diagnosis in 12.6% of the cases (79 probands), and other untiered SNVs (9.4%) and CNVs/SVs or short tandem repeats (STRs) (5.8%). The HL panels were applied to 89% of the cohort (2025 probands) through the standard Genomics England pipeline. 48% of the diagnoses (300) were identified in green genes in the applied HL panels, corresponding to a 13% diagnostic yield across the entire cohort. Genetic diagnoses were identified in 273 different genes. The most common causative variants were found in *USH2A*, accounting for 9% of the total diagnoses. Other frequently diagnosed genes included *GJB2* (5.4%), *COL1A1* (3%), *MYO15A* (2.4%), *MYO7A* (2.1%), *WFS1* (2.1%), *SOX10* (1.9%), *STRC* (1.6%), *TMPRSS3* (1.4%), and *CDH23, MYO6* and *TECTA*, each contributing to 1.3% of the diagnoses. The full list of genetic diagnoses is shown in <u>Supplementary Table 2</u>. Almost half of the causative variants were predicted loss-of-function and 37.1% were missense variants. A smaller proportion was attributed to splicing variants (8.5%), CNV/SV/STR (4.2%) and synonymous, intronic/regulatory and mitochondrial variants (each contributing less than 0.5%). Of solved cases, autosomal dominant (47.2%) and autosomal recessive (47%) modes of inheritance were seen in almost equal frequency. X-linked dominant (3%) or recessive (2.2%) patterns were less commonly observed. An additional 0.5% accounted for uniparental disomy and mitochondrial inheritance. Out of the 367 families recruited with a minimum family structure of three or more members, 117 diagnosed variants were identified as *de novo* (32%). In 81% of cases, the diagnosis was considered actionable and demonstrated clinical utility in 24.6% of instances by informing reproductive choices (15.7%), facilitating additional surveillance for patients or their relatives (7.2%), eliciting changes in medication (0.6%), qualifying for clinical trial participation (0.3%), or other outcomes (0.8%).

A summary of findings for the HED cohort can be found in Figure 1B and <u>Table 2: Diagnostic overview</u>. The diagnostic rate within the cohort is 27.3%, spanning 68 different genes. *GJB2* (MIM #121011) was the most common, followed by *USH2A* (MIM #608400) and *MYO15A* (MIM #602666). These three genes accounted for approximately 30% of the diagnoses. The HL panels were applied to 97.5% of the cohort (755 probands) through the standard Genomics England pipeline. 86.7% of the diagnoses (183) were identified in green genes in the applied HL panels, corresponding to a 23.6% diagnostic yield across the entire cohort. An autosomal recessive mode of inheritance was seen in a larger proportion of diagnoses in this group (63%). There was also a small rise in consanguinity by +0.7%. The full list of genetic diagnoses is shown in <u>Supplementary Table 2</u>.

### Exomiser performance

Exomiser [29, 30] is a phenotype-driven variant prioritisation tool designed to annotate, filter, and prioritise likely causative variants for rare Mendelian diseases.. A retrospective analysis of the 625 diagnoses found this cohort demonstrated that Exomiser successfully identified 597 of these (95.5%), with the causative variant ranked within the top 5 in 89.3% of instances (Figure 2). This performance likely underestimates Exomiser’s true diagnostic potential, as 23 of the missed diagnosed variants (3.7%) were undetectable due to issues such as the absence of the variant or it being flagged as low quality in the input VCF file, or due to technical limitations such as undetectable variations including STR or regions characterised by poor mapping such as *STRC*. Furthermore, 29 diagnoses (4.6%) were identified using settings that accounted for incomplete penetrance.

**Figure 2:**
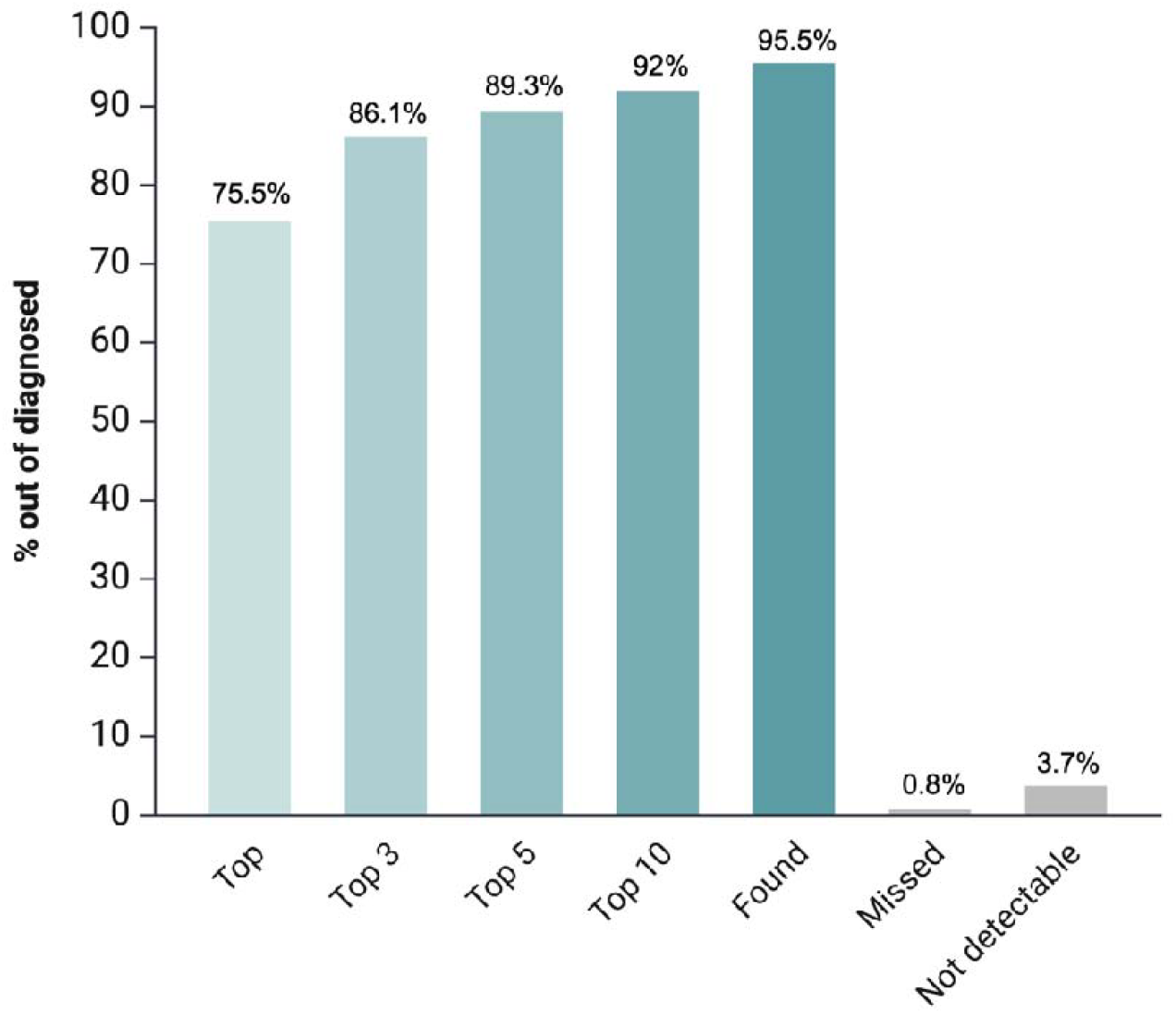
Exomiser performances in ranking the diagnosed variants within the HL cohort. The Exomiser software successfully identified 597 out of 625 diagnoses (95.5%), with 89.3% of these diagnoses ranked within the top 5.

### Assessment of variation of diagnostic rates and prevalent genes after HL stratification

To investigate whether diagnostic rates differed by hearing loss sub-types, we stratified the cohort by auditory phenotype. This revealed some significant differences, as illustrated in Figure 3. In the congenital HL subgroup, the diagnostic rate was higher than in both the childhood-onset and adult-onset groups. However, this difference was only statistically significant for the comparison with adult-onset HL (33.2% vs 16.7% respectively; p-value=0.017, CI=1.15-5.98, OR=2.49). The diagnostic rate was also significantly higher in individuals with bilateral HL (27%) compared to those with unilateral HL (11.1%), as shown in Figure 3 (p-value=7.61e-5, CI=1.64-5.70, OR=2.95). Within the unilateral subgroup, there were 14 diagnoses spanning 13 different genes, predominantly linked to syndromic forms of HL. Notably, probands with diagnoses in *ALG3, CHD7, MICU1, PTPN11*, and *SHANK3* exhibited microcephaly. Additionally, diagnosed genes such as *COL1A1, COL1A2, FGFR1, KAT6, HUWE1*, and *SOX10* are also associated with skeletal or facial development abnormalities

**Figure 3:**
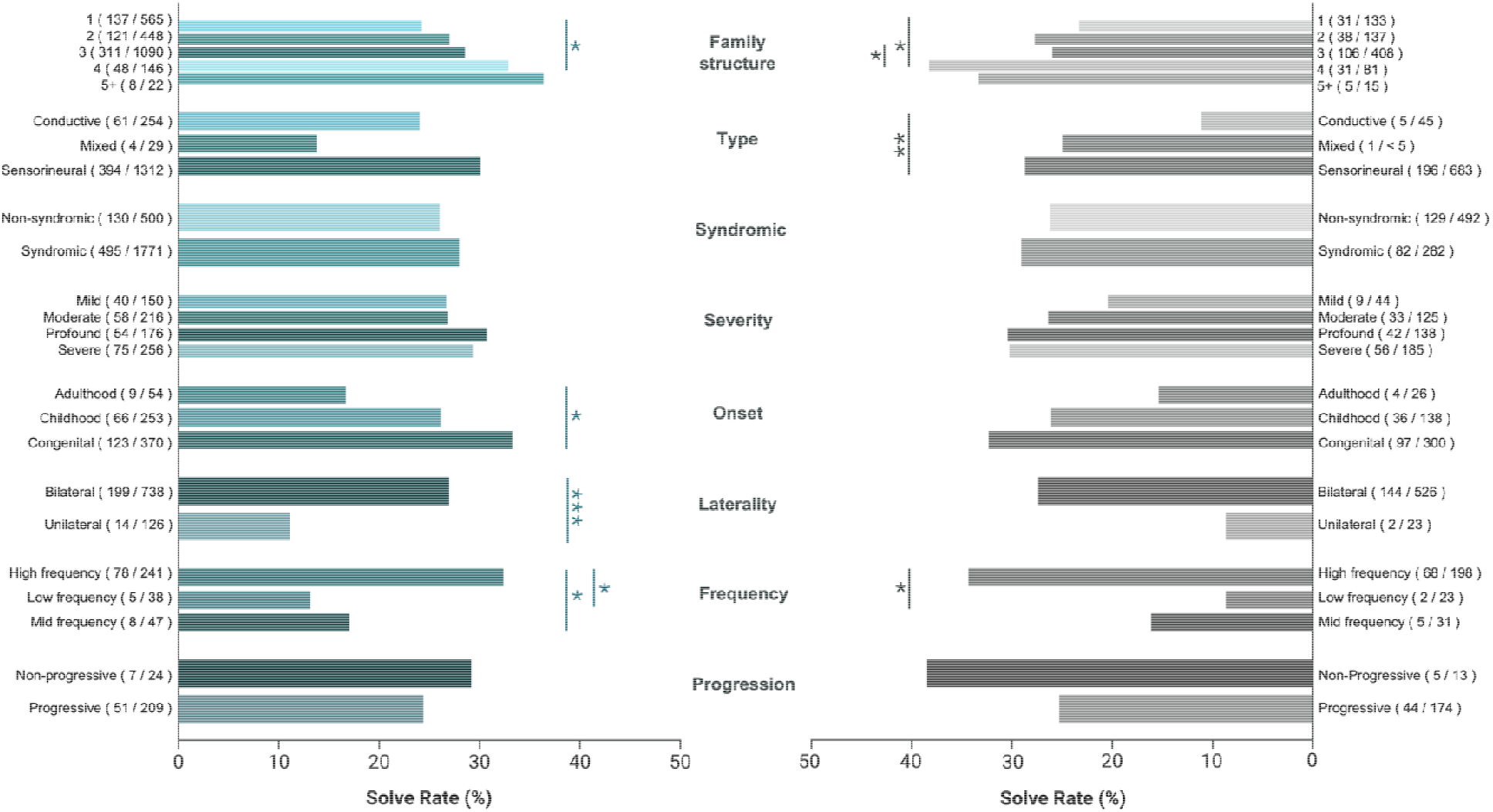
Diagnostic rate within the HL and HED cohort stratified by type, severity, onset, progression, laterality, and frequency of their HL. The ^*^ indicates p values < 0.05 after two-sided Fisher’s exact test, ^**^ p values <0.01 and ^***^ p values <0.001. To ensure compliance with data protection policies, any frequency counts lower than 5 associated with phenotypic information have been redacted and are represented as ‘<5’ and the minimum possible solve rate corresponding to the lowest redacted count is shown.

HL can also vary in the way it affects threshold detection at different frequencies generating different audiogram shapes from flat to sloping at either high or low frequencies because of the tonotopic organisation of the cochlea. The diagnostic rate within the high-frequency HL subgroup (32.4%) exhibited a large and significant increase compared to both the mid-frequency HL group (17%) (p-value=0.037, CI=1.10-6.04, OR=2.33) and the low-frequency HL group (13.2%) (p-value=0.021, CI=1.16-10.73, OR=3.16).

Stratifying into different types of HL revealed that while not statistically significant, the sensorineural HL subset demonstrated the highest diagnostic yield at 30%, encompassing 394 diagnoses out of 1312 probands, followed by conductive (24%) and mixed subset (13.8%) (Figure 3). A gene ontology analysis reveals significant enrichment of diagnosed genes within the conductive HL subset in several categories: chromosome organisation (GO:0051276; FDR adjusted p-value = 5.4e-4), chromatin organisation (GO:0006325; FDR adjusted p-value = 3.9e-4), as well as positive regulation of gene expression (GO:0010628; FDR adjusted p-value = 3.4e-4), transcription (GO:0006355; FDR adjusted p-value = 1.38E-04), and sensory perception of sound (GO:0007605; FDR adjusted p-value = 4.4e-8).

However, the diagnostic rate does not appear to vary significantly based on the severity, progression and syndromic/non-syndromic type of HL. A slight increase in diagnostic rates is observed in cases of profound (30.7%) and severe (29.3%) HL compared to moderate (26.9%) and mild (26.7%) HL; however, this difference was not statistically significant. Although the diagnostic rate was higher in cases reported as non-progressive (29.2%) compared to progressive (24.4%), this difference was also not statistically significant. The diagnostic rates between syndromic and non-syndromic HL were similar, 28% and 26%, respectively; this difference was also not statistically significant. As expected, the diagnostic rate is positively correlated to family structure, particularly the number of recruited individuals within families, where the % solve rate progressively increases from 24.2% in singletons to 32.9% in families with 4 individuals (p-value=0.044, CI=0.43-0.99, OR=0.65) and 36.4% in families with five or more individuals.

Applying a similar stratification approach to the HED cohort reveals similar trends and an additional significant statistical difference in the diagnostic rate among patients diagnosed with sensorineural HL (28.7%) compared to those with conductive HL (11.1%) (p-value=0.0093, CI=1.24-10.59, OR=3.22), as illustrated in Figure 3. Diagnostic rates and statistics from Figure 3 are also available in <u>Supplementary Table 3</u> and <u>Supplementary Table 4</u>.

### Diagnostic rates and prevalent genes by combination of phenotypic abnormalities

The upset diagram presented in Figure 4 illustrates the various combinations of phenotypic abnormalities identified within the cohort, delineating non-syndromic HL and different types of syndromic HL. The bar plot visually represents the number of individuals in each subset, while the line graph highlights the diagnostic yield within each specific subset of patients. Among the non-syndromic cases (n = 500), all HPO terms belonged to the “abnormality of the ear” category. The diagnostic rate within this subset was 26% (130 families), similar to the overall cohort. A full list of diagnoses by phenotypic abnormalities combination is available in <u>Supplementary Table 5</u>. The diagnoses in this subgroup involved genes that are well-established causes of non-syndromic HL, either exclusively or in both syndromic and non-syndromic forms. Examples of such genes include *GJB2* (accounting for 16.9% of the diagnoses in this subgroup), *MYO15A* (7.7%), *USH2A* (6.2%), *STRC* (5.4%), *TMPRSS3* (5.4%), *SLC26A4* and *TECTA* (each contributing to 4.6%), *MYO6* and *TMC1* (each contributing to 3.8%), followed by *CDH23, MYO7A, SOX10, GATA3, LOXHD1, MITF, OTOF, EYA4, GSDME* and *OTOG* each contributing to <3.8%. To ensure compliance with data protection policies, percentages corresponding to counts lower than five have been redacted.

**Figure 4:**
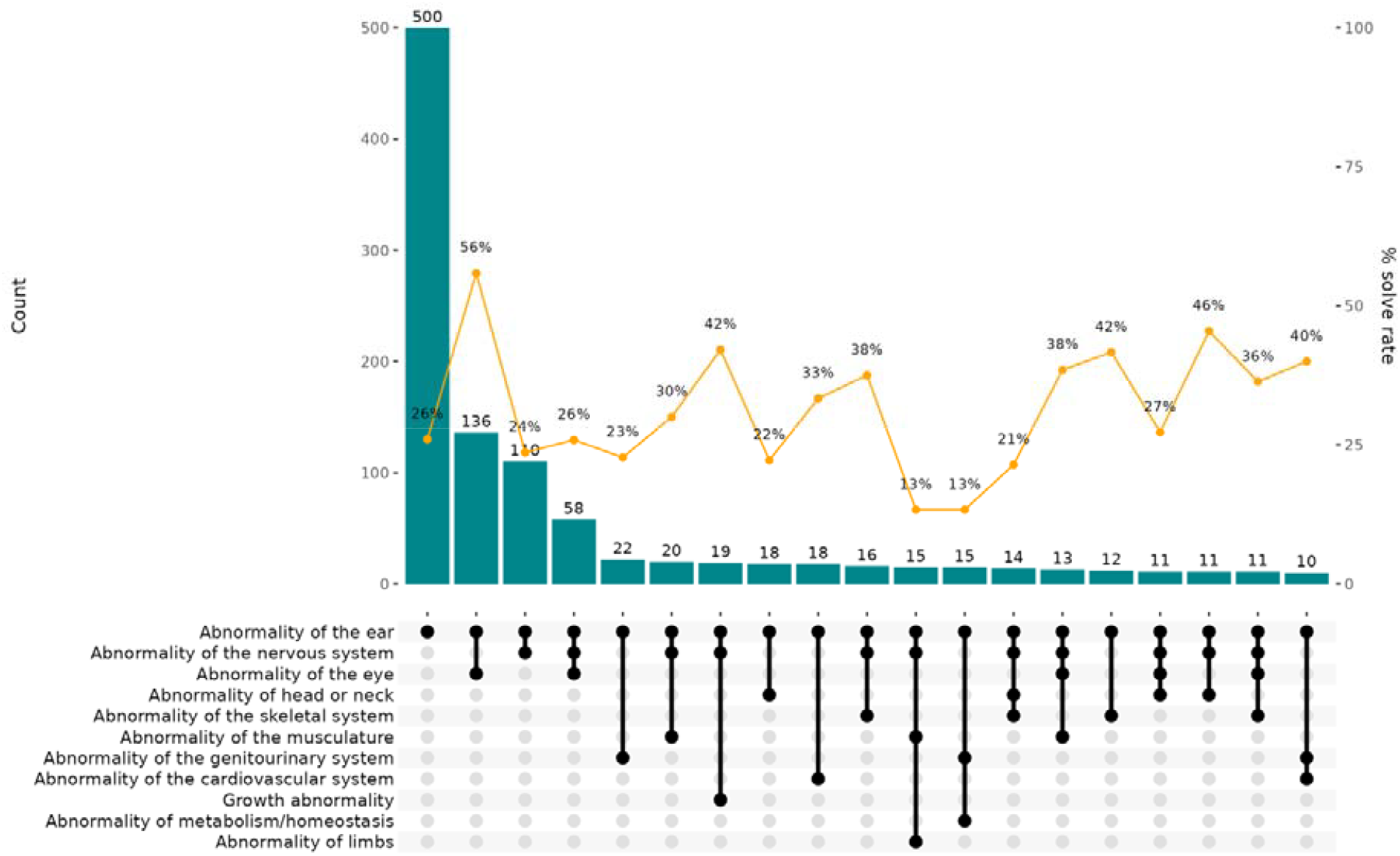
Upset diagram of phenotypic abnormalities within the HL cohort. The plot shows the combination matrix in grey, the relative number of patients within each subset as a bar plot and the relative diagnostic yield (%) as a line graph.

The most frequent syndromic combination of phenotypic abnormalities involved patients with both ear and eye abnormalities. A higher diagnostic yield of 56% (76/136) was reported in individuals with this phenotypic combination. The majority of these were genes associated with Usher syndrome: *USH2A* (accounting for 48.7% of the diagnoses in this subgroup), *MYO7A* (10.5%), followed by *CDH23, CLRN1, USH1C and ADGRV1* (each contributing to <6.6%). Another common gene represented in this subset was *WFS1* (9.2%), associated with Wolfram syndrome. Other rare causes identified include *PRPS1* associated with Arts syndrome and Charcot-Marie-Tooth disease, *ABHD12* associated with polyneuropathy, HL, ataxia, retinitis pigmentosa, and cataract, *COL11A1* associated with Stickler syndrome, *CYP1B1*associated with anterior segment dysgenesis and *GUCY2D* associated with Leber congenital amaurosis and cone-rod dystrophy. Gene ontology analysis revealed a statistically significant enrichment of genes associated with stereocilia cellular components (GO:0032420; p-value = 2.6e-16) within this subgroup.

The next most frequent phenotypic combination involved patients with both ear and nervous system abnormalities. Among the 110 patients in this subset, a diagnosis was made in 26 cases, resulting in a diagnostic yield of 24%. Interestingly, this subset’s most diagnosed genes were *GJB2* (19.2%) and *MYO6* (<19.2%), typically associated with non-syndromic HL. Among the other diagnoses were neurological phenotypes where HL is a well-recognised feature, including *ATP1A3* associated with *CAPOS* syndrome, *FOXC1* associated with Axenfeld-Rieger syndrome, *THOC6* associated with Beaulieu-Boycott-Innes syndrome and *SOX10* which is associated with Waardenburg syndrome. Additionally, there were genes with a neurodevelopmental phenotype where HL has been a rare or occasionally reported feature. Examples of these genes include *EIF2B5* associated with leukoencephalopathy with vanishing white matter, *KAT6A* associated with Arboleda-Tham syndrome, *SETD5* associated with intellectual developmental disorder and *WDR45* associated with neurodegeneration with brain iron accumulation, among others.

A combination of ear, nervous system, and eye was associated with a diagnostic yield of 26%. This included genes such as *OPA1* associated with Behr syndrome and both syndromic and non-syndromic forms of optic atrophy, *ATP1A3* associated with CAPOS syndrome, *POLG* associated with ophthalmoplegia, sensorimotor polyneuropathy, ataxia, and deafness, *PRPS1* associated with Charcot-Marie-Tooth disease, and *LRP2* associated with Donnai-Barrow syndrome, among others.

## Discussion

This research aims to elucidate the genomic architecture of hearing loss in UK patients recruited to the 100kGP. This encompasses not only patients with a predominant HL presentation, which is the primary focus of previously reported HL cohorts, but also those with any hearing-related phenotype, including those seen as part of a more complex presentation. The analysis presented in this study is notable for the inclusion of a particularly large cohort: 2762 individuals affected by either syndromic or non-syndromic HL and 2726 unaffected relatives belonging to 2271 distinct rare disease families recruited to the 100kG. A subset of patients recruited under the disease group “hearing and ear disorders” constitute the HED cohort, for whom HL was considered the predominant clinical phenotype at the time of recruitment. This cohort includes 1067 affected individuals and 972 unaffected relatives, belonging to 774 distinct rare disease families recruited to the 100kG. The overall diagnostic rate achieved in the HL cohort is 27.5% (625/ 2271), and 27.3% (211/ 774) in the HED cohort. This is a very conservative estimate of the diagnostic rate, which includes only fully diagnosed patients and partially diagnosed individuals whose HL is attributed to a variant in a gene marked as green in the HL PanelApp panels. Therefore, it excludes any partially diagnosed individuals whose diagnosed gene is not a green gene in the HL panel, as well as any candidates awaiting confirmation or labelled as VUS.

This diagnostic rate appears to be lower compared with some other HL cohorts described in the literature, often reporting a diagnostic rate between 20% and 50% [10-20]. However, these are often based on panel-based analyses. Several additional factors likely contribute to this difference, including variations in the inclusion/exclusion criteria used across different projects. It is worth noting that the recruitment for the 100kGP allowed for the inclusion of patients with HL who may not strictly meet the criteria of being congenital and/or bilateral. Additionally, a recommendation that participants should undergo pre-screening for known prevalent HL genes during recruitment, such as *GJB2, GJB6*, and *SLC26A4*, undoubtedly impacted the overall diagnostic rates.

Variants in *GJB2, USH2A*, and MYO15A were among the top four diagnoses in the HL and HED cohort. *COL1A1*-related diseases were the third most common diagnosis in the HL cohort. *COL1A1* is associated with a range of conditions, most commonly Osteogenesis Imperfecta, where HL is a frequently reported feature. However, genetic testing was likely to be requested because of the predominant skeletal rather than hearing phenotype.

The top 4 diagnoses accounted for approximately 20% of the total, with diagnoses made in 273 different genes overall; by comparison, only 68 different genes were identified in the HED cohort. This demonstrates the genomic complexity of syndromic HL and that many such conditions are not covered by standard HL panels, which are designed with a focus on non-syndromic presentations.

The size of this HL cohort provides statistical power to allow comparison in diagnostic rates among hearing subcategories. Figure 3 shows a statistically significant increase in the diagnostic rate among patients with high-frequency HL (32.4%) compared to mid-frequency HL (17%) and low-frequency HL (13.2%). This was also seen in bilateral HL (27%) compared to unilateral HL (11.1%) and congenital HL (33.2%) compared to adult-onset HL (16.7%). This difference may reflect the fact that unilateral and adult-onset HL frequently can also stem from environmental or non-genetic causes, such as ageing, noise exposure, infections, trauma, or acquired anomalies in the auditory system’s structure or function. This information helps clinicians understand when genetic testing is more likely to be informative.

Within the HED cohort, statistically significant differences in diagnostic rates were seen between the sensorineural (28.7%) and conductive HL (11.1%) subgroups. This may be partially explained by genes with high prevalence, such as *GJB2, STRC, USH2A*, being typically associated with sensorineural HL. In addition, many of the diagnoses associated with conductive HL have a syndromic presentation and were recruited outside the HED subgroup. Examples of such diagnoses identified in this context include *ANKRD11* associated with KBG syndrome (MIM #148050), *CREBBP* associated with Rubinstein-Taybi syndrome (MIM #180849), *MN1* associated with CEBALID syndrome (MIM #618774) and others. Otitis media or structural changes seen in syndromic presentations may be relevant to the context of conductive HL. Furthermore, such genes are not typically included in standard HL gene panels. Consequently, diagnoses might be missed unless a syndromic diagnosis and the need for broader genetic testing are considered. An interesting observation relates to the statistically significant enrichment of genes associated with chromosome (GO:0051276; FDR adjusted p-value = 5.4e-4) and chromatin organisation (GO:0006325; FDR adjusted p-value = 3.9e-4) in individuals with conductive HL, including *DDX3X, CHD2, KDM5C, ATRX, ACTB, MECOM, POGZ, ARID1B, CREBBP, NSD1, EYA1, SETD5* and *TP63*. Many of these genes would not be included on a standard HL genetic panel. This reinforces the need to think about the most appropriate strategy for genomic investigation in individuals presenting with conductive HL, particularly when associated with additional syndromic features.

A unique aspect of this work was the use of HPO terms to allow interrogation across combinations of phenotypic abnormalities. Firstly, among the subgroup with the phenotype restricted only to abnormalities of the ear, approximately 40% of the diagnoses were in genes that can be associated with additional syndromic features. It is documented that a group of conditions known as ‘non-syndromic mimic’ presents as non-syndromic but might have under-recognised additional clinical features or features that develop later in life [36]. An example of this is *USH2A*, which often presents as non-syndromic HL with later-onset retinal problems. Frequently observed diagnoses in this group include Usher, Pendred, Waardenburg syndromes and *GATA3*-associated HDR (hypoparathyroidism, sensorineural deafness and renal dysplasia). There were also rare cases of DOORS and Noonan syndromes, both of which would be expected to have additional features to HL. The lack of non-HL HPO terms in this group might represent an under-recognition of these features or a phenotype that has not yet developed at the time of recruitment. This reinforces the need for pre-test counselling in anticipation of possible syndromic diagnosis and potential management implications.

Analysis of the group of patients reporting ear and eye abnormalities reveals a very high diagnostic rate (56%), predominantly accounted for by Usher syndrome genes and *WFS1* (Wolfram syndrome). Notably, *USH2A* accounts for 48.7% of the diagnoses in this subgroup. Genes including *USH2A, MYO7A, CDH23, CLRN1, USH1C* and *ADGRV1* are known to be required for the development, maintenance and function of hair cell bundles and stereocilia. This is also evident in the gene ontology analysis, which indicates a statistically significant enrichment in genes associated with stereocilia cellular components (GO:0032420; p-value = 2.6e-16) within this ear-eye subgroup.

The standard 100kGP analysis pipeline, which includes the use of selected gene panels and the review by the clinical scientists of Tier 1 and Tier 2 coding SNV or indel variants, identified the likely genetic diagnosis in 72% of the total confirmed diagnoses reported here. Further curation resulted in diagnoses for an additional 174 families.

This approach included the use of Exomiser across the whole cohort, highlighting a useful role for this software as a diagnostic adjunct in automated pipeline analysis by ranking the diagnostic genomic variant(s) in the top 5 in 89.3% of cases. However, Exomiser will not be able to detect genetic variation that is not present in the VCF file used as input therefore, 3.7% of missed diagnoses were due to factors such as the absence or flagging of the variant as low quality, undetectable variations like STRs or genomic regions characterised by not well covered by short-read sequencing. One example is the low detection rate of *STRC* variants by the standard 100kGP analysis. Biallelic variants, including both SNV and CNV, in *STRC* are one of the most common causes of non-syndromic HL, with an estimated prevalence of approximately 4-10% among non-syndromic HL patients who test negative for the *GJB2* gene [37-39]. However, the detection rate for *STRC* within the HL cohort was lower than expected, at 1.6%. The interpretation of sequence data from the *STRC* gene is challenging due to the presence of a pseudogene that shares 99.6% homology with the functional *STRC* gene. As a result, a significant proportion of short reads may map to both genomic regions, thereby reducing coverage ratios and the ability to accurately call CNVs in that region. Similar issues may also impact the mapping quality of other genes associated with HL that have high sequence similarity with pseudogenes in the vicinity. For instance, the *OTOA* gene exhibits high sequence similarity and approximately 99% homology in the exon 20-28 region, with its pseudogene located 820 kilobases downstream [40, 41]. To address these challenges, the use of long-read sequencing has demonstrated the ability to effectively resolve the *STRC* gene from its pseudogene, although it doesn’t detect all small CNVs yet. Consequently, long-read sequencing and techniques like droplet digital PCR (ddPCR) and long-range PCR (LR-PCR) have been more recently recommended for improved detection and characterisation of these challenging genomic regions [40].

It is also necessary to acknowledge the limitations in our current knowledge in interpreting the pathogenicity of variants outside the exons, especially regarding the role of non-coding variations, enhancer and regulatory regions, as well as situations involving the contribution of multiple genes (oligogenic or polygenic) to the disease. These methodological advancements hold the potential to significantly increase the diagnostic yield of the 100kGP over time. However, it is important to acknowledge that despite these improvements, there will likely remain a subset of HL patients who may never receive a definitive diagnosis. This includes individuals whose HL is partly or wholly caused by environmental or non-genetic factors, as well as those with multifactorial causes, presenting inherent challenges in achieving a conclusive diagnosis. Although there are great advantages of working with such a large cohort of patients recruited across the UK, it is also important to acknowledge the challenge of dealing with missing outcome data, incomplete entry of HPO terms and limited clinical descriptions of patients’ phenotypes. This has undoubtedly affected the overall diagnostic rate. The use of HPO terms in this dataset offers several advantages but also limitations. One significant advantage is that it provides a standardised framework for characterising patients’ clinical features, allowing seamless data integration and comparison across different recruiting centres. It also facilitates the use of phenotype-based prioritisation tools such as Exomiser. However, it is crucial to acknowledge certain limitations in using this ontology within the 100kGP, particularly with respect to HL, where diagnoses are more complex than some other diseases. One notable concern is the potential for incomplete or missing information in the selection of HPO terms by the recruiting clinician. The selection process may be influenced by the clinician’s field of expertise, which may not comprehensively capture all the intricacies of a patient’s phenotype. There is a high percentage of missing data regarding the specific auditory phenotypes in some of the syndromic patients. For example, information regarding the type of HL was missing in 29.8% of the HL cohort, similarly on severity (64.9%), onset (70.2%), progression (89.7%), laterality (62%), and frequency (85.6%). Auditory phenotypes were more comprehensively completed in patients recruited to the HED cohort, where we have a reduction of missing data between 14 and 33% across subsets. Additionally, it is important to consider that HPO annotations represent a static snapshot of the phenotypic presentation at the time of recruitment. They may not encompass the dynamic nature of the disease, and some of the patients’ clinical features may evolve over time. Nevertheless, it is worth noting that efforts have been made to automatically integrate HPO and ICD-10 terms directly from electronic health records, which may partially address some of these issues and improve the completeness and accuracy of HPO-based analyses going forward.

Furthermore, despite the completion of the recruitment phase, the analysis of the 100kGP is still ongoing. It is anticipated that the diagnostic rate will continue to increase in the future as newly identified candidate genes emerge, novel tools become available, and further evidence potentially leads to the reclassification of some variants currently classified as VUS. In this regard, there is a clear need for the development of functional studies or assays to help resolve this large proportion of VUS and elucidate the functional implications of these variants. Additionally, periodic updates of PanelApp genes and re-analysis of the data [42], integration of gene expression and multi-omics analysis, and the use of novel tools may provide further insights into potential functional consequences and mechanisms of pathogenicity in the future.

## Conclusions

The permissive inclusion criteria allowed the analysis of patients with any hearing or ear-related phenotypes. This allowed insight into the true range of genetic pathologies impacting the auditory system beyond phenotypes typically included in HL cohorts. Unlike previous studies that predominantly focused on non-syndromic presentations or common HL syndromes, this study encompassed a wider array of phenotypes, shedding light on the comprehensive genetic heterogeneity associated with HL and reinforcing the need to consider broader testing outside the HL panel, especially for individuals with a syndromic presentation where conductive HL is a feature. Notably, Exomiser emerged as a valuable and effective tool for identifying genetic diagnoses related to HL, highlighting its utility in identifying the correct diagnoses in diverse phenotypic contexts. Furthermore, leveraging the statistical power of this large dataset allowed quantification of the likelihood of genetic diagnosis with specific phenotypic combinations and identified positive predictors of a genetic diagnosis.

Consequently, this study offers valuable insights into the genomic and phenotypic architecture of HL in the UK, and the data generated as part of this work will contribute to advancing scientific understanding, improving diagnostic approaches, informing clinical care and supporting future research efforts in the field.

## Data Availability

The source data is available through the Genomics England Research Environment subject to successful application. Results of the data analysis are presented within Figures and Supplementary Data in the manuscript. To ensure compliance with data protection policies, any frequency counts lower than 5 associated with phenotypic information have been redacted and are represented as “<5”; this is indicated in the footnote below whenever it occurs.

## Acknowledgements

We dedicate this work to the late Professor Maria Bitner-Glindzicz, whose recruitment of hundreds of patients with hearing loss and great dedication to their care were the true inspiration for this research. This research was made possible through access to data in the National Genomic Research Library, which is managed by Genomics England Limited (a wholly owned company of the Department of Health and Social Care). The National Genomic Research Library holds data provided by patients and collected by the NHS as part of their care and data collected as part of their participation in research. The National Genomic Research Library is funded by the National Institute for Health Research and NHS England. The Wellcome Trust, Cancer Research UK and the Medical Research Council have also funded research infrastructure. The analysis was supported by PhD funding from the National Institute for Health and Care Research University College London Hospitals Biomedical Research Centre Deafness And Hearing Problems Theme (Grant Ref BRC392/HD/AS/110390) and a grant from the NIH, National Institute of Child Health and Human Development 1R01HD103805-01. The work was also funded by Biomedical Research Centre Deafness And Hearing Problems Theme (Grant Ref BRC872/HD/OF/110390 to EC) and a grant from the Medical Research Council (MC_UP_1503/2 and MR/X004597/1 to MRB). We extend our gratitude to the Genomics England Diagnostic Discovery Hub and Susan Walker, Head of Translational Genomics, for their significant contributions to identifying diagnoses in this patient cohort and the Rare Disease cohort as a whole. We also thank the NHS England Genomics Unit, the NHS Genomic Laboratory Hubs (NHS GLHs), all the NHS recruiting clinicians, the research community, and the patients and families recruited to the 100,000 Genomes Project for their invaluable contributions.

## Conflict of interests

The authors declare the following competing interests: D.S. was seconded to and received salary from Genomics England, a wholly owned Department of Health and Social Care company, from 2016-2018. All other authors have no competing interests. The Authors confirm the independence of researchers from funders and that all authors had full access to all of the data in the study and can take responsibility for the integrity of the data and the accuracy of the data analysis.

## Data Sharing

The source data is available through the Genomics England Research Environment subject to successful application. Results of the data analysis are presented within Figures and Supplementary Data in the manuscript. To ensure compliance with data protection policies, any frequency counts lower than 5 associated with phenotypic information have been redacted and are represented as ‘<5’; this is indicated in the footnote below whenever it occurs.

## Summary box

### What is already known on this topic

- Although mutations in numerous genes can cause human hearing loss, their respective contributions to the frequency and type of inherited deafness are largely unknown.
- The opportunity to perform Whole Genome Sequencing (WGS) and to catalogue all variants present in a patient’s DNA as part of the 100,000 Genomes Project (100kGP), and correlate these with the progression of their specific hearing phenotype across a large UK clinical population, has potential to bridge the gap between current academic knowledge, clinical practice and patient benefit.
- Hearing loss studies previously reported usually focus on non-syndromic cohorts only.

### What this study adds

- This study reports on a large cohort consisting of 2271 rare disease families with varying degrees of hearing impairment recruited to the 100kGP. This group comprises 5488 individuals, 2762 affected individuals, and 2726 unaffected relatives.
- A subcohort of patients recruited under the “hearing and ear disorders” category, for whom the hearing loss likely manifests as the predominant clinical phenotype, includes 774 families. This group comprises 2039 individuals, 1067 affected individuals, and 972 unaffected relatives.
- The study’s permissive inclusion criteria including any proband having at least one HPO term descendant of the “Hearing impairment” (HP:0000365), allow for a broad range of auditory phenotypes to be represented, improving understanding of both syndromic and non-syndromic hearing loss.
- Disease stratification using HPO terms reveals a different likelihood of diagnosis and prevalence of genes per subtype and combination of phenotypic abnormalities.
- Exomiser proves to be a valuable and effective tool for identifying HL diagnoses that have not been solved through routine diagnostic pathways.
- Whole genome sequencing offers a more advanced and refined method for detecting HL diagnoses, that may not be identified by standard diagnostic techniques and targeted disease panels

## Notes

### Author Declarations

The 100,000 Genomes Project (100kGP) is a UK Government project launched in 2013 and coordinated by Genomics England, a wholly owned company of the Department of Health and Social Care. The project aims to apply whole-genome sequencing to the study of rare diseases, cancers, and infections in a national healthcare setting. After ethical approval (National Research Ethics approvals 14/EE/1112 and 13/EE/032), NHS rare disease patients (probands and family members) were recruited to the 100kGP Rare Disease programme (pilot or main) through one of 13 Genomic Medicine Services (GMCs) around the UK. All participants provided written informed consent. Project RR140 was registered as part of the Hearing and Sight Genomics England Clinical Interpretation Partnership (GeCIP), reviewed and accepted by Genomics England through an Access Review Committee and the Ethics Advisory Committee.

