## Supplementary Table 1 for "The genomic landscape of syndromic and non-syndromic hearing loss within the 100,000 Genomes Project cohort"

| HPO term | HPO definition |
|---|---|
| HP:0000365 | Hearing impairment |
| HP:0000405 | Conductive hearing impairment |
| HP:0000410 | Mixed hearing impairment |
| HP:0008513 | Bilateral conductive hearing impairment |
| HP:0008591 | Congenital conductive hearing impairment |
| HP:0008598 | Mild conductive hearing impairment |
| HP:0008607 | Progressive conductive hearing impairment |
| HP:0012716 | Moderate conductive hearing impairment |
| HP:0012717 | Severe conductive hearing impairment |
| HP:0040119 | Unilateral conductive hearing impairment |
| HP:0000407 | Sensorineural hearing impairment |
| HP:0000408 | Progressive sensorineural hearing impairment |
| HP:0001757 | High-frequency sensorineural hearing impairment |
| HP:0008504 | Moderate sensorineural hearing impairment |
| HP:0008527 | Congenital sensorineural hearing impairment |
| HP:0008573 | Low-frequency sensorineural hearing impairment |
| HP:0008587 | Mild neurosensory hearing impairment |
| HP:0008615 | Adult onset sensorineural hearing impairment |
| HP:0008619 | Bilateral sensorineural hearing impairment |
| HP:0008625 | Severe sensorineural hearing impairment |
| HP:0011474 | Childhood onset sensorineural hearing impairment |
| HP:0000399 | Prelingual sensorineural hearing impairment |
| HP:0008596 | Postlingual sensorineural hearing impairment |
| HP:0008610 | Infantile sensorineural hearing impairment |
| HP:0011476 | Profound sensorineural hearing impairment |
| HP:0040113 | Old-aged sensorineural hearing impairment |
| HP:0011975 | Aminoglycoside-induced hearing loss |
| HP:0001730 | Progressive hearing impairment |
| HP:0005101 | High-frequency hearing impairment |
| HP:0008542 | Low-frequency hearing loss |
| HP:0009900 | Unilateral deafness |
| HP:0012712 | Mild hearing impairment |
| HP:0012713 | Moderate hearing impairment |
| HP:0012714 | Severe hearing impairment |
| HP:0012715 | Profound hearing impairment |
| HP:0012779 | Transient hearing impairment |
| HP:0012781 | Mid-frequency hearing loss |

**Supplementary Table 1: HPO terms used to define the cohort of 2,271 HL probands included in this study**
This table lists the 37 Human Phenotype Ontology (HPO) terms descendant of "Hearing Impairment" (HP:0000365) that were used to define the cohort of 2,271 HL probands included in this study
