## Supplementary Table 2 for "The genomic landscape of syndromic and non-syndromic hearing loss within the 100,000 Genomes Project cohort"

| Case solved summary | Diagnosed gene | Diagnosed variant | Exomiser rank | Exomiser MOI | Tier | Assembly | HED |
|---|---|---|---|---|---|---|---|
| diagnosed | AARS1 | 16:70302259:C:T | 1 | AD | NA | b37 | N |
| diagnosed | ABHD12 | 20:25290213:T:C | 1 | AR | 3 | b37 | N |
| diagnosed | ABHD14A-ACY1 | 3:51985205:A:G\|3:51987601:G:A | 7 | AR | 3 | b38 | N |
| diagnosed | ACTB | 7:5528589:A:G | 1 | AD | 1 | b38 | Y |
| diagnosed | ACTB | 7:5527849:C:T | 1 | AD | 1 | b38 | N |
| diagnosed | ACTG1 | 17:81511362:G:A | 4 | AD | 1 | b38 | N |
| diagnosed | ACVRL1 | 12:51913720:G:T | 1 | AD | 1 | b38 | N |
| diagnosed | ADGRV1 | 5:90690946:C:T\|5:90855813:CAT:C | 1 | AR | 1 | b38 | N |
| diagnosed | ADGRV1 | 5:90644818:TG:T\|5:90778583:C:T | 1 | AR | 1 | b38 | N |
| diagnosed | ADGRV1 | 5:90755015:C:T | 1 | AR | 1 | b38 | Y |
| partially ( HL explained ) | ADGRV1 | 5:90642887:G:A | 1 | AR | 2 | b38 | N |
| diagnosed | ADGRV1 | 5:90658078:CAG:C | 179 | AR | 3 | b38 | Y |
| partially ( HL explained ) | ADGRV1\|NR2F1 | CNV | 2 | AD | - | | N |
| diagnosed | ADNP | 20:49520426:T:TTC | 1 | AD | 1 | b37 | N |
| diagnosed | ADNP | 20:49508932:C:CT | 1 | AD | 1 | b37 | N |
| diagnosed | AHDC1 | 1:27548104:G:A | 4 | AD | 1 | b38 | N |
| diagnosed | AHDC1 | 1:27548302:G:A | 4 | AD | 1 | b38 | N |
| diagnosed | AIFM1 | X:130137134:A:G | 1 | XR | 2 | b38 | N |
| diagnosed | AIFM1 | X:130139869:C:T | 1 | XR | 2 | b38 | N |
| diagnosed | ALG3 | 3:184248776:G:A | 1 | AR | 2 | b38 | N |
| diagnosed | ALMS1 | 2:73453093:AC:A\|2:73572702:CAG:C | 1 | AR | 1 | b38 | N |
| diagnosed | ALMS1 | 2:73449036:G:GT | 1 | AR | 1 | b38 | N |
| diagnosed | ALMS1 | 2:73718084:C:T\|CNV | 2 | AR | - | b37 | N |
| diagnosed | ANKRD11 | 16:89345770:G:A | 1 | AD | 1 | b37 | N |
| diagnosed | ANKRD11 | 16:89284140:TTTTC:T | 1 | AD | 1 | b38 | N |
| diagnosed | ANKRD11 | 16:89285643:G:C | 1 | AD | 1 | b38 | N |
| diagnosed | ANKRD11 | 16:89281907:TG:T | 1 | AD | 1 | b38 | N |
| diagnosed | ANKRD11 | 16:89280028:C:CG | 1 | AD | 1 | b38 | N |
| diagnosed | ANKRD11 | 16:89279749:C:CG | 1 | AD | 1 | b38 | N |
| diagnosed | ANKRD11 | 16:89284772:CTTCAGCGA:C | 1 | AD | 1 | b38 | N |
| diagnosed | ANKRD11\|ARX | 16:89351956:TGGCCTTGTGCTTGA:T\|X:25033774:G:C | 1 | XD | NA | b37 | N |
| diagnosed | ARID1B | 6:157198907:G:A | 1 | AD | 1 | b38 | N |
| diagnosed | ARID1B | 6:156779328:C:T | 1 | AD | 1 | b38 | N |
| diagnosed | ARID1B | 6:157511345:G:A | 1 | AD | 1 | b37 | N |
| diagnosed | ARL6\|IMPG2 | 3:97791825:A:G\|3:101231117:G:A | 1 | AR | 2 | b38 | N |
| diagnosed | ASXL2 | 2:25743885:C:CA | 1 | AD | 1 | b38 | N |
| diagnosed | ASXL3 | 18:33743212:C:T | 1 | AD | 3 | b38 | N |
| diagnosed | ATAD3A | 1:1459703:C:CG | 8 | AR | NA | b37 | N |
| diagnosed | ATP1A3 | 19:41970275:C:T | 1 | AD | 1 | b38 | N |
| diagnosed | ATP1A3 | 19:41970275:C:T | 1 | AD | 2 | b38 | N |
| diagnosed | ATP1A3 | 19:41970275:C:T | 1 | AD | 2 | b38 | N |
| diagnosed | ATP1A3 | 19:42474427:C:T | 1 | AD | 1 | b37 | N |
| diagnosed | ATP6V1B2 | 8:20220382:C:T | 2 | AD | 3 | b38 | Y |
| diagnosed | ATP6V1B2 | 8:20220382:C:G | 1 | AD | 1 | b38 | Y |
| diagnosed | ATP6V1B2 | 8:20077893:C:T | NA | AD | NA | b37 | N |
| diagnosed | ATRX | X:77574322:C:T | 1 | XR | 2 | b38 | N |
| diagnosed | ATXN3 | STR | NA | AD | - | | N |
| diagnosed | BBS1 | 11:66526181:T:G | 2 | AR | 2 | b38 | N |
| diagnosed | BBS1 | 11:66293652:T:G | 1 | AR | 2 | b37 | N |
| diagnosed | BCAP31 | X:153702971:G:A | 1 | XR | 1 | b38 | N |
| diagnosed | BEST1 | 11:61724904:G:A | 2 | AD | 2 | b37 | N |
| diagnosed | BSND | 1:55007173:GC:G | 1 | AR | 1 | b38 | N |
| diagnosed | BSND | 1:54999209:G:A\|1:55007173:GC:G | 1 | AR | 1 | b38 | Y |
| partially ( HL explained ) | BTK\|TIMM8A | X:101348603:T:TGCTGCAACTGCGGGTCC | 1 | XR | NA | b38 | N |
| diagnosed | CACNA1A | 19:13261526:C:T | 1 | AD | 1 | b38 | N |
| diagnosed | CACNA1A | 19:13224773:C:T | 1 | AD | 1 | b38 | N |
| diagnosed | CACNA1A | 19:13262820:CT:C | 1 | AD | 1 | b38 | N |
| diagnosed | CACNA1G | 17:50599722:C:T | 1 | AD | NA | b38 | N |
| diagnosed | CAMTA1 | 1:7806025:AT:A | 6 | AD | 1 | b37 | N |
| diagnosed | CCNO | 5:55231577:G:A\|5:55233275-55233279:DUP | 1 | AR | - | b38 | N |
| diagnosed | CDH23 | 10:73539073:G:A\|10:73571115:C:A | 1 | AR | 2 | b37 | Y |
| diagnosed | CDH23 | 10:71803362:A:G | 3 | AR | 2 | b38 | Y |

| | | | | | | | |
|---|---|---|---|---|---|---|---|
| diagnosed | CDH23 | 10:71712781:G:C\|10:71740853:G:A | 1 | AR | 2 | b38 | Y |
| diagnosed | CDH23 | 10:71712781:G:C\|10:71808007:G:A | 5 | AR | 2 | b38 | N |
| diagnosed | CDH23 | 10:73537991:G:C\|10:73565607:C:CAAGGATG | 1 | AR | 1 | b37 | Y |
| diagnosed | CDH23 | 10:71709206:C:A | 1 | AR | 2 | b38 | Y |
| diagnosed | CDH23 | 10:73442332:A:T | 1 | AR | 2 | b37 | Y |
| diagnosed | CDH23 | 10:71646579:G:A\|10:71809915:CACA:C | 1 | AR | 3 | b38 | N |
| diagnosed | CDK13 | 7:40047840:G:C | 2 | AD | 1 | b38 | N |
| diagnosed | CDK5RAP2 | 9:120439621:T:TC | 2 | AR | 1 | b38 | N |
| diagnosed | CHAMP1 | 13:114324384:CTG:C | 9 | AD | 1 | b38 | N |
| diagnosed | CHD2 | 15:93002269:CAAAG:C | 1 | AD | 1 | b38 | N |
| diagnosed | CHD7 | 8:60741663:AC:A | 1 | AD | 1 | b38 | N |
| diagnosed | CHD7 | 8:60801526:A:G | 1 | AD | 1 | b38 | Y |
| diagnosed | CHD7 | 8:60852027:AGT:A | 1 | AD | 1 | b38 | N |
| diagnosed | CHD7 | 8:60837010:C:T | 1 | AD | 1 | b38 | N |
| partially ( HL explained ) | CHD7 | 8:60852584:G:A | 1 | AD | 1 | b38 | N |
| diagnosed | CHD7 | 8:60842051:G:T | 1 | AD | 1 | b38 | N |
| diagnosed | CLIC5 | 6:45949316:TC:T | 1 | AR | 1 | b38 | Y |
| diagnosed | CLRN1 | 3:150972557:C:A | 1 | AR | 1 | b38 | N |
| diagnosed | CLRN1 | 3:150972557:C:A\|3:150972591:A:C | 3 | AR | NA | b38 | Y |
| diagnosed | CLRN1 | 3:150972488:C:T\|3:150972591:A:C | 4 | AR | 2 | b38 | N |
| diagnosed | CLRN1 | 3:150972642:C:A\|CNV | 1 | AR | - | b38 | N |
| diagnosed | CLTC | 17:59644366:G:T | 1 | AD | 1 | b38 | N |
| diagnosed | CLTC\|RERE | 17:59685644:C:T\|1:8360224:C:CCT | 1 | AD | 1 | b38 | N |
| diagnosed | COCH | 14:30878834:G:A | 1 | AD | 2 | b38 | Y |
| partially ( HL explained ) | COL11A1 | 1:103449693:C:A | 1 | AD | 3 | b37 | N |
| partially ( HL explained ) | COL11A1 | 1:103435770:C:T | 1 | AD | 1 | b37 | N |
| diagnosed | COL11A1 | 1:103474073:CT:C | 1 | AD | 1 | b37 | N |
| diagnosed | COL11A1 | CNV | 2 | AD | - | b37 | N |
| diagnosed | COL11A1 | 1:103008517:CT:C | 8 | AD | 3 | b38 | N |
| partially ( HL explained ) | COL11A2 | 6:33167305:G:A | 1 | AD | 1 | b38 | N |
| partially ( HL explained ) | COL11A2 | 6:33178166:C:A | 3 | AD | 3 | b38 | N |
| diagnosed | COL11A2 | 6:33166512:C:T | 1 | AD | 3 | b38 | N |
| diagnosed | COL11A2 | 6:33170555:A:T | 1 | AD | NA | b38 | Y |
| diagnosed | COL1A1 | 17:50189395:A:AG | 1 | AD | 1 | b38 | N |
| diagnosed | COL1A1 | 17:50197770:G:A | 1 | AD | 1 | b38 | N |
| diagnosed | COL1A1 | 17:50199310:AG:A | 1 | AD | 1 | b38 | N |
| diagnosed | COL1A1 | 17:50195457:G:A | 1 | AD | 1 | b38 | N |
| diagnosed | COL1A1 | 17:50199310:AG:A | 1 | AD | 1 | b38 | N |
| diagnosed | COL1A1 | 17:50188765:G:A | 1 | AD | 1 | b38 | N |
| diagnosed | COL1A1 | 17:50195231:C:G | 1 | AD | 1 | b38 | N |
| diagnosed | COL1A1 | 17:50195641:G:A | 1 | AD | 1 | b38 | N |
| diagnosed | COL1A1 | 17:50189406:T:TC | 1 | AD | 1 | b38 | N |
| diagnosed | COL1A1 | 17:50190036:CA:C | 1 | AD | 1 | b38 | N |
| diagnosed | COL1A1 | 17:50188643:G:T | 3 | AD | 2 | b38 | N |
| diagnosed | COL1A1 | 17:50189201:T:A | 113 | AD | NA | b38 | N |
| diagnosed | COL1A1 | 17:50197045:C:T | 1 | AD | 2 | b38 | N |
| diagnosed | COL1A1 | 17:50195961:CG:C | 1 | AD | 1 | b38 | N |
| diagnosed | COL1A1 | 17:50197234:C:T | 1 | AD | 1 | b38 | N |
| diagnosed | COL1A1 | 17:50188115:ACAGG:A | 1 | AD | 1 | b38 | N |
| diagnosed | COL1A1 | 17:48273002:G:A | 1 | AD | 1 | b37 | N |
| diagnosed | COL1A1 | 17:50199416:AC:A | NA | AD | NA | b38 | N |
| diagnosed | COL1A1 | 17:50199255:CG:C | NA | AD | NA | b38 | N |
| diagnosed | COL1A2 | 7:94410501:G:C | 1 | AD | 2 | b38 | N |
| diagnosed | COL1A2 | 7:94409795:G:A | 1 | AD | 2 | b38 | N |
| diagnosed | COL1A2 | 7:94409795:G:A | 1 | AD | 2 | b38 | N |
| diagnosed | COL1A2 | 7:94034006:G:T | 1 | AD | 2 | b37 | N |
| diagnosed | COL2A1 | 12:47982910:C:A | 4 | AD | 2 | b38 | N |
| diagnosed | COL2A1 | 12:47995872:T:A | 1 | AD | 1 | b38 | N |
| diagnosed | COL2A1 | 12:47980960:A:ACGTTCACC | 1 | AD | 1 | b38 | N |
| diagnosed | COL2A1 | 12:47985772:C:T | 1 | AD | 1 | b38 | N |
| diagnosed | COL4A3 | 2:227297727:G:C | 1 | AD | 2 | b38 | N |
| diagnosed | COL4A4 | 2:227056044:C:T | 1 | AD | 2 | b38 | N |
| diagnosed | COL4A4 | 2:227088678:C:T | 1 | AR | 2 | b38 | N |

| | | | | | | | |
|---|---|---|---|---|---|---|---|
| diagnosed | COL4A5 | X:108626299:G:A | 1 | XD | 2 | b38 | N |
| diagnosed | COL4A5 | X:108601921:G:T | 1 | XR | 2 | b38 | N |
| partially ( HL explained ) | COL4A5 | X:108578120:G:C | 3 | XD | 1 | b38 | N |
| diagnosed | COL4A5 | X:108597386:G:A | 1 | XD | 2 | b38 | N |
| partially ( HL explained ) | COL4A5 | X:108598825:G:C | 1 | XD | 2 | b38 | N |
| diagnosed | COL4A5 | X:108603062:G:A | 1 | XD | 1 | b38 | N |
| diagnosed | CREBBP | 16:3749631:C:T | 1 | AD | 1 | b38 | N |
| diagnosed | CREBBP | 16:3729478:GCTGGAT:G | NA | AD | NA | b38 | N |
| diagnosed | CSNK2A1 | 20:492282:T:C | 1 | AD | 1 | b38 | N |
| diagnosed | CYP1B1 | 2:38071008:TC:T | 1 | AR | 1 | b38 | N |
| diagnosed | DDX3X | X:41343801:C:T | 1 | XD | NA | b38 | N |
| diagnosed | DDX3X | X:41346541:CAT:C | 1 | XD | 1 | b38 | N |
| diagnosed | DHX30 | 3:47849725:C:T | 83 | AD | 2 | b38 | N |
| diagnosed | DNAAF4 | NA | 1 | AR | 3 | b38 | N |
| diagnosed | DNAH11 | 7:21900087:G:T\|7:21901076:C:T | 10 | AR | 1 | b38 | N |
| diagnosed | DNAH5 | 5:13769605:C:T\|5:13830872:C:CGTGAT | 1 | AR | 1 | b37 | N |
| diagnosed | DNAH5 | 5:13701316:T:TA\|CNV | 1 | AR | - | b38 | N |
| diagnosed | DNAI2 | 17:74309345:G:A | 1 | AR | 1 | b38 | N |
| diagnosed | DNAJC3 | 13:95790877:GAGAA:G | 1 | AR | 3 | b38 | N |
| diagnosed | DNMT1\|PRKCG | 19:10159683:G:A\|19:53889654:A:G | 1 | AD | 3 | b38 | N |
| diagnosed | DONSON\|SON | 21:33554981:CAGTT:C | 1 | AD | 1 | b38 | N |
| diagnosed | DPF2 | 11:65346043:GACTGTGGCCGCTCAGGT:G | 1 | AD | 1 | b38 | N |
| diagnosed | DPF2 | 11:65348877:G:A | 1 | AD | 3 | b38 | N |
| diagnosed | DSP | 6:7575697:GGATA:G | 2 | AD | NA | b37 | N |
| diagnosed | DYRK1A | 21:37505590:G:A | 2 | AD | 1 | b38 | N |
| diagnosed | EBF3 | 10:129848455:GTAATC:G | 2 | AD | 1 | b38 | N |
| diagnosed | EBF3\|GJB2 | 10:129867246:G:A\|13:20189212:G:A\|13:20189511:C:T | 2 | AR | 1 | b38 | N |
| diagnosed | ECHS1 | 10:133364716:A:AT | 1 | AD | 3 | b38 | N |
| diagnosed | EFTUD2 | 17:44851720:C:T | 1 | AD | 1 | b38 | Y |
| diagnosed | EFTUD2 | CNV | 7 | AD | - | | N |
| diagnosed | EGR2 | 10:62813572:C:G | 24 | AD | 2 | b38 | N |
| diagnosed | EIF2B5 | 3:183854445:G:A\|3:183854453:GA:G | 1 | AR | 1 | b37 | N |
| diagnosed | EIF3F | 11:7994466:T:G | 3 | AR | 3 | b38 | N |
| diagnosed | EP300 | 22:41168600:G:GT | 1 | AD | 1 | b38 | N |
| diagnosed | EPS8L2 | 11:723327:A:AC | 1 | AR | 3 | b37 | Y |
| diagnosed | ERBB2\|PGAP3 | 17:39687876:T:TCAGAGCGCCCCCAGAG | 1 | AR | 1 | b38 | N |
| diagnosed | ERCC8 | 5:60194144:G:A\|5:60200619:C:T | 1 | AR | 3 | b37 | N |
| diagnosed | ESPN | 1:6460100:G:A | 1 | AR | 1 | b38 | Y |
| diagnosed | EXT1 | 8:118110564:T:TA | 1 | AD | 1 | b38 | N |
| diagnosed | EYA1 | 8:71321761:CT:C | 1 | AD | 1 | b38 | Y |
| diagnosed | EYA1 | 8:71269739:C:G | 1 | AD | 1 | b38 | N |
| diagnosed | EYA1 | 8:71215455:G:T | 1 | AD | 1 | b38 | N |
| diagnosed | EYA1 | 8:72211338:TG:T | 1 | AD | 1 | b37 | N |
| diagnosed | EYA1 | 8:71216687:C:T | 5 | AD | 3 | b38 | N |
| diagnosed | EYA1 | CNV | NA | AD | - | | Y |
| partially ( HL explained ) | EYA4 | 6:133512780:G:A | 4 | AD | 1 | b38 | N |
| partially ( HL explained ) | EYA4 | 6:133525156:A:T | 2 | AD | 1 | b38 | N |
| diagnosed | EYA4 | 6:133464858:G:A | 26 | AD | 2 | b38 | Y |
| diagnosed | EYA4 | 6:133464858:G:A | 9 | AD | 2 | b38 | Y |
| diagnosed | FBP1 | 9:94603557:C:T | 3 | AR | 3 | b38 | Y |
| diagnosed | FGF3 | 11:69625259:G:C\|11:69631128:C:T | 1 | AR | 2 | b37 | Y |
| diagnosed | FGFR1\|IL17RD | 8:38279354:C:T\|3:57135235:T:C | 1 | AD | NA | b37 | N |
| diagnosed | FGFR2 | 10:123247561:C:G | 209 | AD | 3 | b37 | Y |
| partially ( HL explained ) | FGFR3 | 4:1805644:C:A | 1 | AD | 2 | b38 | Y |
| diagnosed | FGFR3 | 4:1804392:G:A | 1 | AD | 2 | b38 | N |
| partially ( HL explained ) | FGFR3 | 4:1801844:C:G | 138 | AD | NA | b38 | N |
| diagnosed | FOXC1 | 6:1611217:G:T | 1 | AD | 3 | b38 | Y |
| diagnosed | FOXC1 | CNV | 8 | AD | - | | Y |
| diagnosed | FRAS1 | 4:78333404:GT:G\|4:78387367:C:G | 3 | AR | 1 | b38 | N |
| diagnosed | FXN | STR | NA | AR | - | b37 | Y |
| diagnosed | FZD2 | 17:44558988:G:A | 1 | AD | 3 | b38 | N |
| diagnosed | G6PC3\|NSD1 | 17:44071095:C:T\|5:177267565:G:T | 1 | AR | 2 | b38 | N |
| diagnosed | GATA3 | 10:8100566:GAA:G | 1 | AD | NA | b37 | Y |

| | | | | | | | |
|---|---|---|---|---|---|---|---|
| diagnosed | GATA3 | 10:8069511:T:A | 1 | AD | 1 | b38 | Y |
| diagnosed | GATA3 | 10:8058764:TC:T | 1 | AD | 1 | b38 | Y |
| diagnosed | GATA3 | CNV | NA | AD | - | | N |
| diagnosed | GATA3 | CNV | 2 | AD | - | | N |
| diagnosed | GDAP1 | 8:74360184:C:T | 1 | AD | 2 | b38 | N |
| diagnosed | GJB2 | 13:20189546:AC:A | 1 | AR | 1 | b38 | Y |
| diagnosed | GJB2 | 13:20189255:CCCCTTGATGAACTT:C\|13:20189546:AC:A | 1 | AR | 1 | b38 | Y |
| diagnosed | GJB2 | 13:20189358:C:T | 1 | AD | 2 | b38 | Y |
| diagnosed | GJB2 | 13:20189511:C:T | 1 | AR | 1 | b38 | Y |
| diagnosed | GJB2 | 13:20188953:AAT:A\|13:20189546:AC:A | 1 | AR | 1 | b38 | N |
| diagnosed | GJB2 | 13:20189481:A:G\|13:20189546:AC:A | 1 | AR | NA | b38 | Y |
| partially ( HL explained ) | GJB2 | 13:20763497:C:T | 2 | AD | 2 | b37 | N |
| partially ( HL explained ) | GJB2 | 13:20189299:C:T\|13:20189481:A:G | 1 | AR | NA | b38 | Y |
| diagnosed | GJB2 | 13:20189546:AC:A | 3 | AR | 1 | b38 | Y |
| partially ( HL explained ) | GJB2 | 13:20188997:C:T\|13:20192783:C:A | 3 | AR | 2 | b38 | Y |
| diagnosed | GJB2 | 13:20189546:AC:A | 1 | AR | 1 | b38 | Y |
| diagnosed | GJB2 | 13:20189388:T:C\|13:20189546:AC:A | 3 | AR | 1 | b38 | Y |
| diagnosed | GJB2 | 13:20189546:AC:A | 1 | AR | 1 | b38 | Y |
| diagnosed | GJB2 | 13:20189546:AC:A | 1 | AR | 1 | b38 | Y |
| diagnosed | GJB2 | 13:20189546:AC:A | 1 | AR | 1 | b38 | Y |
| partially ( HL explained ) | GJB2 | 13:20189546:AC:A | 1 | AR | 1 | b38 | N |
| diagnosed | GJB2 | 13:20189155:G:A\|13:20189511:C:T | 1 | AR | 1 | b38 | Y |
| diagnosed | GJB2 | 13:20189414:CA:C\|13:20189481:A:G | 1 | AR | NA | b38 | Y |
| diagnosed | GJB2 | 13:20189351:C:T | 2 | AR | 1 | b38 | Y |
| diagnosed | GJB2 | 13:20189351:C:T\|13:20189546:AC:A | 1 | AR | 1 | b38 | Y |
| diagnosed | GJB2 | 13:20189351:C:T | 1 | AD | 1 | b38 | Y |
| partially ( HL explained ) | GJB2 | 13:20189481:A:G | 1 | AR | NA | b38 | N |
| diagnosed | GJB2 | 13:20189546:AC:A | 1 | AR | 1 | b38 | N |
| diagnosed | GJB2 | 13:20189450:C:T\|13:20189546:AC:A | 1 | AR | 1 | b38 | Y |
| diagnosed | GJB2 | 13:20189291:G:GT\|13:20189546:AC:A | 1 | AR | 1 | b38 | Y |
| partially ( HL explained ) | GJB2 | 13:20763685:AC:A | 1 | AR | NA | b37 | Y |
| diagnosed | GJB2 | 13:20763685:AC:A | 1 | AR | NA | b37 | Y |
| diagnosed | GJB2 | 13:20189546:AC:A | NA | AR | NA | b38 | Y |
| diagnosed | GJB2 | 13:20189481:A:G | 1 | AR | NA | b38 | Y |
| diagnosed | GJB2 | 13:20189546:AC:A | NA | AR | NA | b38 | Y |
| partially ( HL explained ) | GJB2 | 13:20189481:A:G\|CNV | 1 | AR | - | b38 | N |
| diagnosed | GJB2 | 13:20189481:A:G | 1 | AR | NA | b38 | Y |
| diagnosed | GJB2 | 13:20189546:AC:A | 2 | AR | NA | b38 | Y |
| diagnosed | GLA | X:101398942:T:C | 1 | XR | 2 | b38 | N |
| diagnosed | GNAS | 20:58909703:C:A | 1 | AD | 3 | b38 | N |
| diagnosed | GNAS | 20:58903687:A:G | 1 | AD | 1 | b38 | N |
| diagnosed | GNB1 | 1:1815799:G:GCCC | 1 | AD | 1 | b38 | N |
| diagnosed | GRXCR1 | 4:42963075:C:T | 1 | AR | 1 | b38 | Y |
| diagnosed | GSDME | 7:24706179:C:T | 10 | AD | NA | b38 | Y |
| diagnosed | GSDME | 7:24706378:T:C | 2 | AD | NA | b38 | Y |
| diagnosed | GUCY2D | 17:7905914:T:C\|17:7918713:C:A | NA | AR | NA | b37 | Y |
| diagnosed | H3-3A | 1:226071433:C:T | 5 | AD | 3 | b38 | N |
| diagnosed | H4C9 | 6:27139430:G:T | 27 | AD | 3 | b38 | N |
| diagnosed | HDAC8 | X:72572110:CT:C | 1 | XD | 1 | b38 | N |
| diagnosed | HDAC8 | X:72572706:A:C | 1 | XD | 1 | b38 | N |
| diagnosed | HIKESHI | 11:86055766:A:G | 1 | AR | NA | b37 | N |
| diagnosed | HSD17B4 | 5:119478985:G:GCGGGATCA\|5:119493821:G:A | 1 | AR | 1 | b38 | N |
| partially ( HL explained ) | HSD17B4 | 5:119452618:A:G | 1 | AR | 2 | b38 | Y |
| diagnosed | HUWE1 | X:53536657:A:C | 1 | XR | 2 | b38 | N |
| diagnosed | HUWE1 | X:53536576:T:C | 1 | XD | 1 | b38 | N |
| diagnosed | INF2 | 14:105169535:T:C | 1 | AD | 2 | b37 | N |
| diagnosed | KANSL1 | 17:46171269:GC:G | 1 | AD | 1 | b38 | N |
| diagnosed | KANSL1 | 17:46170854:C:G | 1 | AD | 1 | b38 | N |
| diagnosed | KARS1 | 16:75669683:T:A\|16:75669880:G:A | 5 | AR | NA | b37 | N |
| diagnosed | KAT6A | 8:41978773:CAT:C | 1 | AD | 1 | b38 | N |
| diagnosed | KAT6A | 8:41949334:C:T | 1 | AD | 1 | b38 | N |
| diagnosed | KAT6B | 10:76744953:C:CA | 1 | AD | 1 | b37 | N |
| diagnosed | KCNE1 | NA | NA | AR | NA | | Y |

| | | | | | | | |
|---|---|---|---|---|---|---|---|
| diagnosed | KCNJ10 | 1:160041976:A:T\|1:160042363:G:C | 1 | AR | 2 | b38 | N |
| diagnosed | KCNQ1 | 11:2549213:TA:T | 1 | AR | 1 | b37 | Y |
| diagnosed | KCNQ4 | 1:40784388:T:TTCGTCTACC | 1 | AD | 2 | b38 | N |
| diagnosed | KCNQ4 | 1:40820180:G:A | 4 | AD | 2 | b38 | N |
| diagnosed | KCNQ4 | 1:40819912:C:A | 1 | AD | 2 | b38 | Y |
| partially ( HL explained ) | KCNQ4 | 1:40818642:T:G | 1 | AD | 3 | b38 | Y |
| diagnosed | KCNQ4 | 1:41285569:A:C | 1 | AD | 2 | b37 | Y |
| diagnosed | KDM5C | X:53193559:A:AG | 1 | XR | NA | b38 | N |
| diagnosed | KDM6A | X:45061375:C:T | 1 | XD | 3 | b38 | N |
| diagnosed | KIF11 | 10:92606263:A:G | 10 | AD | 3 | b38 | N |
| diagnosed | KIF1A | 2:241737138:C:T | 1 | AD | 2 | b37 | N |
| diagnosed | KIT | 4:54736508:C:T | 1 | AD | 2 | b38 | N |
| diagnosed | KMT2A | 11:118352591:C:T | 1 | AD | 2 | b37 | N |
| diagnosed | KMT2C | 7:152315249:ATAAATCCAGGCGT:A | 2 | AD | 1 | b38 | N |
| diagnosed | KMT2C | 7:152247986:G:T | 10 | AD | 3 | b38 | N |
| diagnosed | KMT2E | 7:105112579:A:AT | 11 | AD | 3 | b38 | N |
| diagnosed | LDB3 | 10:86687163:G:A | 1 | AD | 2 | b38 | N |
| diagnosed | LHFPL5 | 6:35773532:T:TG\|6:35773578:T:C | 1 | AR | 1 | b37 | Y |
| partially ( HL explained ) | LHFPL5 | 6:35806065:G:A | 12 | AR | 1 | b38 | N |
| diagnosed | LMNB2 | 19:2434305:C:T | 1 | AD | 3 | b38 | N |
| diagnosed | LOXHD1 | 18:46529200:TC:T | 1 | AR | 1 | b38 | Y |
| partially ( HL explained ) | LOXHD1 | 18:46529227:G:A | 2 | AR | 1 | b38 | N |
| partially ( HL explained ) | LOXHD1 | 18:46524707:CCTT:C\|18:46656978:TACAGGGCCAGG:T | 1 | AR | 1 | b38 | Y |
| diagnosed | LOXHD1 | 18:46529227:G:A | 1 | AR | 1 | b38 | Y |
| diagnosed | LOXHD1 | 18:46505875:GC:G | 1 | AR | 1 | b38 | Y |
| diagnosed | LOXHD1 | 18:46579688:G:A | 2 | AR | 2 | b38 | Y |
| diagnosed | LRP2 | 2:170059380:G:A\|2:170060657:T:A | 1 | AR | 1 | b37 | N |
| diagnosed | LRP2 | 2:170060532:A:C\|2:169917941-170339028:DEL | 995 | AR | - | b37 | N |
| diagnosed | LRP2 | 2:169206877:A:C\|2:169280361:C:T | 5 | AR | 1 | b38 | N |
| diagnosed | LZTR1 | 22:20991684:G:A | 12 | AD | 2 | b38 | N |
| diagnosed | MAF | 16:79633612:G:C | 1 | AD | NA | b37 | N |
| diagnosed | MAGEL2 | 15:23644699:T:TG | 1 | AD | 3 | b38 | N |
| diagnosed | MAGEL2 | 15:23645837:G:A | 1 | AD | 1 | b38 | N |
| diagnosed | MAN2B1 | 19:12649146:A:G\|19:12657475:GGC:G | 1 | AR | 1 | b38 | N |
| diagnosed | MAN2B1 | 19:12657482:G:T\|19:12663807:C:T | 1 | AR | 1 | b38 | N |
| diagnosed | MARVELD2 | 5:69432677:T:C | 1 | AR | 1 | b38 | Y |
| diagnosed | MECOM | 3:169100903:G:A | 1 | AD | 3 | b38 | N |
| diagnosed | MECOM | 3:169095235:A:G | 1 | AD | 3 | b38 | N |
| diagnosed | MED12 | X:71128673:GAAGATCTGAT:- | NA | XD | NA | b38 | N |
| diagnosed | MED13L | 12:115983212:G:GC | 1 | AD | 1 | b38 | N |
| diagnosed | MED13L | 12:116421085:G:A | 1 | AD | 1 | b37 | N |
| diagnosed | MEG3 | CNV | NA | UPD | - | | N |
| diagnosed | MFSD8 | 4:127932983:AC:A\|4:127943940:C:CTA | 1 | AR | 1 | b38 | N |
| diagnosed | MICU1 | 10:72408038:C:G | 10 | AR | 1 | b38 | N |
| diagnosed | MID1 | X:10423069:A:ACCTT | 1 | XR | 1 | b37 | N |
| diagnosed | MIP\|WFS1 | 12:56848037:C:A\|4:6303668:G:A | 2 | AD | NA | b37 | Y |
| diagnosed | MITF | 3:69956479:T:C | 1 | AD | 1 | b38 | Y |
| diagnosed | MITF | 3:69959325:C:T | 1 | AD | 1 | b38 | Y |
| diagnosed | MITF | 3:69964897:G:A | 1 | AD | NA | b38 | Y |
| diagnosed | MITF | 3:69956482:A:G | 1 | AD | 2 | b38 | Y |
| diagnosed | MITF | 3:69959280:C:T | 1 | AD | NA | b38 | Y |
| diagnosed | MME | 3:154834478:AC:A\|3:154862224:C:CA | 2 | AR | 3 | b37 | N |
| diagnosed | MN1 | 22:28195937:G:A | 2 | AD | 3 | b37 | N |
| diagnosed | MN1 | 22:27799196:A:AG | 1 | AD | 3 | b38 | N |
| diagnosed | MORC2 | 22:30949777:C:G | 1 | AD | 3 | b38 | N |
| diagnosed | MORC2 | 22:30958692:G:A | 1 | AD | 1 | b38 | N |
| diagnosed | MORC2 | 22:30958684:C:T | 1 | AD | 3 | b38 | N |
| diagnosed | MORC2 | 22:30941503:G:A | 1 | AD | 1 | b38 | N |
| diagnosed | MORC2 | 22:31337490:G:A | 1 | AD | 3 | b37 | Y |
| diagnosed | MPDZ | 9:13183487:TAAAA:T | 1 | AR | 3 | b38 | N |
| diagnosed | MPV17 | 2:27313058:C:T | 1 | AR | 2 | b38 | N |
| diagnosed | MT-ATP6 | M:8617:A:AT | NA | MT | - | | N |
| diagnosed | MYBPC3\|TMPRSS3 | 11:47370092:C:G\|21:43808545:G:T\|21:43809151:TG:T | 1 | AD | NA | b37 | N |

| | | | | | | | |
|---|---|---|---|---|---|---|---|
| partially ( HL explained ) | MYO15A | 17:18149488:TCAGA:T\|17:18167724:G:A | 1 | AR | 1 | b38 | N |
| diagnosed | MYO15A | 17:18135756:C:T\|17:18155335:C:T | 1 | AR | 1 | b38 | Y |
| partially ( HL explained ) | MYO15A | 17:18023247:TC:T\|17:18052802:TCAGA:T | 1 | AR | 1 | b37 | Y |
| partially ( HL explained ) | MYO15A | 17:18142183:T:G\|17:18150820:TGCCACCTCTCCCCAG:T | 14 | AR | 1 | b38 | Y |
| diagnosed | MYO15A | 17:18057087:A:T\|17:18058028:G:A | 1 | AR | 1 | b37 | Y |
| diagnosed | MYO15A | 17:18119933:TC:T | 1 | AR | 1 | b38 | Y |
| diagnosed | MYO15A | 17:18173904:C:CA | 1 | AR | 1 | b38 | Y |
| diagnosed | MYO15A | 17:18024897:GC:G\|17:18046140:C:T | 1 | AR | 1 | b37 | Y |
| diagnosed | MYO15A | 17:18052173:C:T | 2 | AR | 2 | b37 | Y |
| diagnosed | MYO15A | 17:18142233:GGGCCCGT:G | 1 | AR | 1 | b38 | Y |
| partially ( HL explained ) | MYO15A | 17:18136684:G:A\|17:18143797:G:A | 1 | AR | 1 | b38 | Y |
| diagnosed | MYO15A | 17:18121477:C:T\|17:18151952:G:A | 1 | AR | 1 | b38 | Y |
| partially ( HL explained ) | MYO15A | 17:18178766:C:CA | 1 | AR | 1 | b38 | Y |
| diagnosed | MYO15A | 17:18141116:G:A\|17:18148570:T:A | 1 | AR | 1 | b38 | Y |
| diagnosed | MYO15A | 17:18120457:C:CG\|17:18126457:G:T | 1 | AR | 1 | b38 | Y |
| diagnosed | MYO6 | 6:75881704:C:T | 1 | AD | 1 | b38 | Y |
| partially ( HL explained ) | MYO6 | 6:75879924:A:T | 2 | AD | 1 | b38 | Y |
| diagnosed | MYO6 | 6:75890240:A:ATT | 1 | AD | 1 | b38 | Y |
| diagnosed | MYO6 | 6:75881791:C:T | 1 | AD | 1 | b38 | Y |
| diagnosed | MYO6 | 6:75890237:C:T | 1 | AD | 1 | b38 | Y |
| partially ( HL explained ) | MYO6 | 6:75848436:GT:G | 106 | AD | 1 | b38 | Y |
| diagnosed | MYO6 | 6:75866575:AAGGCTTCATTATC:A | 1 | AR | 3 | b38 | Y |
| partially ( HL explained ) | MYO6 | 6:75886094:G:A | 1 | AD | 2 | b38 | Y |
| diagnosed | MYO7A | 11:77156958:C:T | 1 | AD | 2 | b38 | Y |
| diagnosed | MYO7A | 11:77172750:C:T\|11:77190108:G:A | 1 | AR | 1 | b38 | Y |
| diagnosed | MYO7A | 11:76858842:A:G\|11:76912556:CG:C | 1 | AR | 1 | b37 | N |
| diagnosed | MYO7A | 11:77175465:G:A | 1 | AR | 1 | b38 | N |
| diagnosed | MYO7A | 11:77213858:A:G | 1 | AD | 1 | b38 | N |
| diagnosed | MYO7A | 11:77147796:A:G\|11:77190108:G:A | 1 | AR | 1 | b38 | N |
| diagnosed | MYO7A | 11:77190708:CA:C\|11:77201546:G:A | 1 | AR | 1 | b38 | N |
| diagnosed | MYO7A | 11:77147796:A:G\|11:77208452:ACTT:A | 1 | AR | 1 | b38 | N |
| diagnosed | MYO7A | 11:77161030:A:T | 1 | AR | 1 | b38 | Y |
| diagnosed | MYO7A | 11:77142783:C:A\|11:77205554:T:C | 1 | AR | 1 | b38 | Y |
| diagnosed | MYO7A | 11:77190708:CA:C\|11:77208429:G:A | 1 | AR | 1 | b38 | N |
| diagnosed | MYO7A | 11:76912591:G:A | 1 | AR | 2 | b37 | Y |
| diagnosed | MYO7A | 11:76868036:C:T\|11:76901875:AC:A | 1 | AR | NA | b37 | N |
| diagnosed | MYOT | 5:137870830:C:G | 1 | AD | 2 | b38 | N |
| diagnosed | NDRG1 | 8:133259223:A:C | 26 | AR | 2 | b38 | N |
| diagnosed | NDUFA1\|RNF113A | X:119870921:C:CT | 2 | XR | 3 | b38 | N |
| diagnosed | NDUFAF8 | 17:81239402:C:CCGCCTCCG\|17:81239949:C:T | 6 | AR | NA | b38 | N |
| diagnosed | NDUFV1 | 11:67611972:C:T | 1 | AR | 2 | b38 | N |
| diagnosed | NF1 | 17:31229061:C:T | 2 | AD | 1 | b38 | N |
| diagnosed | NF1 | 17:31349197:CACAGA:C | 1 | AD | 1 | b38 | N |
| diagnosed | NF1 | 17:31338092:C:T | 1 | AD | 1 | b38 | N |
| diagnosed | NF2 | CNV | 1 | AD | - | | N |
| diagnosed | NPC1 | 18:23538564:G:C | 1 | AR | 2 | b38 | N |
| diagnosed | NPC1\|CCDC114 | 18:23534544:C:T\|19:48312078:CT:C | 1 | AR | 3 | b38 | N |
| diagnosed | NSD1 | 5:177280733:T:C | 1 | AD | 1 | b38 | N |
| partially ( HL explained ) | OFD1 | X:13736668:CA:C | 1 | XD | 3 | b38 | N |
| diagnosed | OPA1 | 3:193643992:G:A | 1 | AD | 1 | b38 | N |
| diagnosed | OPA1 | 3:193667167:CAGTT:C | 1 | AD | 1 | b38 | N |
| diagnosed | OPA1 | 3:193618926:ATT:A | 1 | AD | 1 | b38 | N |
| diagnosed | OTOA | 16:21697824:A:T\|CNV | 178 | AR | - | b38 | Y |
| diagnosed | OTOF | 2:26474053:G:A\|2:26475912:A:C | 1 | AR | 1 | b38 | Y |
| diagnosed | OTOF | 2:26473569:T:G\|2:26477210:G:A | 1 | AR | 1 | b38 | Y |
| diagnosed | OTOF | 2:26480883:A:AGC\|2:26483474:AAAG:A | 3 | AR | 1 | b38 | Y |
| diagnosed | OTOG | 11:17569155:G:A\|11:17574818:GGT:G | 1 | AR | 1 | b38 | Y |
| diagnosed | OTOG | 11:17593274:TCC:T\|11:17629257:C:G | 1 | AR | 1 | b38 | Y |
| partially ( HL explained ) | OTOG | 11:17574807:GT:G\|11:17633787:C:T | 1 | AR | 1 | b38 | N |
| diagnosed | OTX2 | CNV | 1 | AD | - | | Y |
| diagnosed | PACS2 | 14:105834449:G:A | 1 | AD | 3 | b37 | N |
| diagnosed | PAX3 | 2:222297158:G:C | 1 | AD | 1 | b38 | N |
| diagnosed | PAX3 | CNV | NA | AD | - | b37 | Y |

| | | | | | | | |
|---|---|---|---|---|---|---|---|
| diagnosed | PAX3 | CNV | 1 | AD | - | | Y |
| diagnosed | PBX1 | CNV | 1 | AD | - | b37 | N |
| diagnosed | PBX1 | 1:164792533:A:AC | NA | AD | NA | b38 | N |
| diagnosed | PCDH15 | 10:54317359:G:T | 1 | AR | 2 | b38 | Y |
| diagnosed | PDHA1 | X:19358940:GGAAGTAA:G | 1 | XD | 3 | b38 | N |
| diagnosed | PDZD7 | 10:101009290:ATCT:A\|10:101011924:C:T | 2 | AR | 1 | b38 | Y |
| diagnosed | PDZD7 | 10:101010781:CT:C\|10:101030053:C:CG | 1 | AR | 1 | b38 | Y |
| diagnosed | PEX1 | 7:92130876:C:T\|7:92157748:A:G | 1 | AR | NA | b37 | N |
| diagnosed | PEX11B | 1:145912332:ACCTC:A | 1 | AR | 1 | b38 | N |
| diagnosed | PHKB | 16:47660731:CT:C\|16:47665952:C:T | 2 | AR | 1 | b38 | N |
| diagnosed | PHYH | 10:13288355:C:A\|10:13288543:T:C | 1 | AR | 1 | b38 | N |
| diagnosed | PHYH | 10:13330355:C:A\|10:13337608:T:C | 1 | AR | 1 | b37 | N |
| diagnosed | PHYH\|SERPING1 | 10:13336579:A:AT\|11:57367850:G:A | 1 | AR | 3 | b37 | N |
| diagnosed | PITX3 | 10:102230638:G:T | 1 | AD | 1 | b38 | N |
| diagnosed | PJVK | 2:179319253:C:T\|2:179320828:C:T | 1 | AR | NA | b37 | Y |
| diagnosed | PKD2 | 4:88008243:CG:C | 1 | AD | 1 | b38 | N |
| diagnosed | PKD2 | 4:88019499:C:T | 1 | AD | 1 | b38 | N |
| diagnosed | PKD2 | 4:88038371:C:T | 1 | AD | 2 | b38 | N |
| diagnosed | PKD2 | 4:88007930:A:AC | NA | AD | NA | b38 | Y |
| diagnosed | PLOD2 | 3:146070772:C:T\|3:146071125:G:A | 1 | AR | 1 | b38 | N |
| diagnosed | PLS1 | 3:142684312:G:A | 1 | AD | 3 | b38 | Y |
| diagnosed | PNPT1 | 2:55910966:C:T | 1 | AR | 2 | b37 | N |
| diagnosed | POGZ | 1:151405606:C:CT | 1 | AD | 1 | b38 | N |
| diagnosed | POLG | 15:89323460:C:G | 1 | AD | 3 | b38 | N |
| diagnosed | POLG | 15:89327201:C:T | 1 | AR | 2 | b38 | N |
| diagnosed | POU3F4 | X:82764226:CA:C | 1 | XR | 1 | b37 | Y |
| diagnosed | PPP1R12A | 12:79798511:A:AT | 1 | AD | 3 | b38 | N |
| diagnosed | PRPS1 | X:107645286:C:T | 1 | XD | 1 | b38 | N |
| diagnosed | PRPS1 | X:107645215:C:A | 1 | XR | 2 | b38 | N |
| diagnosed | PRPS1 | X:106888516:C:T | 1 | XD | 1 | b37 | Y |
| diagnosed | PRRT2 | CNV | 2 | AD | - | | N |
| diagnosed | PRX | 19:40397262:G:A | 2 | AR | 1 | b38 | N |
| diagnosed | PRX | 19:40396564:AG:A\|19:40397250:G:A | 1 | AR | 1 | b38 | N |
| diagnosed | PSMD12 | 17:65337046:C:T | 1 | AD | 1 | b37 | N |
| diagnosed | PSMD12 | 17:67357537:CAG:C | 1 | AD | 1 | b38 | N |
| diagnosed | PTPN11 | 12:112450398:C:T | 1 | AD | 1 | b38 | N |
| diagnosed | PTPN11 | 12:112489105:A:C | 1 | AD | 3 | b38 | Y |
| diagnosed | PTPN11 | 12:112477720:A:G | 1 | AD | 1 | b38 | Y |
| diagnosed | PTPN11 | 12:112446385:A:G | 1 | AD | 2 | b38 | Y |
| diagnosed | PTPN11 | 12:112489084:G:A | 1 | AD | 3 | b38 | N |
| diagnosed | PTPN11 | 12:112477719:A:G | 1 | AD | 2 | b38 | N |
| diagnosed | PTPN11 | 12:112450352:A:C | 1 | AD | 2 | b38 | N |
| partially ( HL explained ) | PTPRQ | 12:80484607:T:C | 2 | AR | 1 | b38 | Y |
| partially ( HL explained ) | PTPRQ | 12:80670365:C:T | 12 | AR | 1 | b38 | N |
| diagnosed | PTRH2 | 17:59697656:C:T | 1 | AR | 3 | b38 | N |
| diagnosed | PUF60 | 8:143818483:C:T | 1 | AD | 1 | b38 | N |
| diagnosed | PURA | 5:140114639:G:T | 37 | AD | 2 | b38 | N |
| diagnosed | PUS7 | 7:105111146:CAG:C | 1 | AR | 3 | b37 | Y |
| diagnosed | RNU4ATAC | 2:121530887:C:T\|2:121530928:A:G | NA | AR | NA | b38 | N |
| diagnosed | RPS19 | 19:42373773:C:G | 1 | AD | 2 | b37 | N |
| diagnosed | RSPH1 | 21:42486463:T:G | 1 | AR | 1 | b38 | N |
| diagnosed | RSPH4A | 6:116617083:C:T | 1 | AR | 1 | b38 | N |
| diagnosed | S1PR2 | 19:10224130:C:CAG | 1 | AR | 3 | b38 | Y |
| diagnosed | SALL1 | 16:51141328:T:TCCAGCTGCTGCTG | 1 | AD | 1 | b38 | Y |
| diagnosed | SALL1 | 16:51175718:T:TTAGCAACC | 1 | AD | 1 | b37 | N |
| diagnosed | SALL1 | 16:51139106:GA:G | 1 | AD | 1 | b38 | N |
| diagnosed | SALL1\|ACADM | 16:51141396:G:A\|1:75761161:A:G | 1 | AD | 1 | b38 | N |
| diagnosed | SCN8A | 12:51699761:T:A | 1 | AD | 1 | b38 | N |
| diagnosed | SERAC1 | 6:158532512:A:AT | 1 | AR | 1 | b37 | N |
| diagnosed | SETD2 | 3:47088172:G:A | 1 | AD | 1 | b38 | N |
| diagnosed | SETD2 | 3:47088172:G:A | 1 | AD | 2 | b38 | N |
| diagnosed | SETD5 | 3:9453738:G:A | 1 | AD | 1 | b38 | N |
| diagnosed | SETD5 | 3:9443400:TTA:T | 1 | AD | 1 | b38 | N |

| | | | | | | | |
|---|---|---|---|---|---|---|---|
| diagnosed | SETD5 | 3:9464580:GA:G | 4 | AD | 1 | b38 | N |
| diagnosed | SF3B4 | 1:149923731:T:C | 1 | AD | 1 | b38 | N |
| diagnosed | SH3TC2 | 5:149042716:G:T | 1 | AD | 3 | b38 | N |
| diagnosed | SHANK3 | 22:50730755:T:TCAGCG | 1 | AD | 1 | b38 | N |
| diagnosed | SIAH1 | 16:48395811:C:G | 1 | AD | 3 | b37 | N |
| diagnosed | SIX1 | 14:60648804:T:C | 2 | AD | 2 | b38 | Y |
| diagnosed | SIX1 | 14:60649134:TC:T | 1 | AD | 1 | b38 | Y |
| partially ( HL explained ) | SIX1 | 14:61115391:T:G | 1 | AD | 1 | b37 | Y |
| diagnosed | SKI | 1:2228825:C:G | 1 | AD | 1 | b38 | N |
| diagnosed | SLC26A2 | 5:149977662:GAAAGT:G\|5:149980851:G:A | 1 | AR | 1 | b38 | N |
| diagnosed | SLC26A4 | 7:107661726:G:C\|7:107674970:G:T | 1 | AR | 2 | b38 | Y |
| diagnosed | SLC26A4 | 7:107674970:G:T\|7:107702038:G:A | 1 | AR | 2 | b38 | Y |
| diagnosed | SLC26A4 | 7:107661637:A:G\|7:107661643:T:C | 1 | AR | NA | b38 | Y |
| diagnosed | SLC26A4 | 7:107675060:T:A | 1 | AR | 2 | b38 | Y |
| diagnosed | SLC26A4 | 7:107674970:G:T | 1 | AR | 2 | b38 | Y |
| diagnosed | SLC26A4 | 7:107672245:G:T\|7:107690125:A:G | 2 | AR | 2 | b38 | Y |
| diagnosed | SLC26A4 | 7:107674970:G:T\|7:107690220:A:C | 1 | AR | 2 | b38 | Y |
| diagnosed | SLC29A3 | 10:71362510:G:T | 1 | AR | 1 | b38 | N |
| diagnosed | SLC52A2 | 8:144359860:T:C\|8:144360408:G:A | 1 | AR | 2 | b38 | N |
| diagnosed | SMAD4 | 18:51078306:A:G | 1 | AD | 1 | b38 | N |
| diagnosed | SMAD4 | 18:51078306:A:G | 1 | AD | 1 | b38 | N |
| diagnosed | SMPX | X:21743819:TG:T | 1 | XR | 1 | b38 | Y |
| diagnosed | SNAP29 | 22:20881100:C:CA | 1 | AR | 1 | b38 | N |
| diagnosed | SOX10 | 22:37978005:C:A | 1 | AD | 1 | b38 | Y |
| diagnosed | SOX10 | 22:37983537:T:G | 1 | AD | 2 | b38 | Y |
| diagnosed | SOX10 | 22:37977943:G:T | 1 | AD | 1 | b38 | Y |
| diagnosed | SOX10 | 22:37983778:C:CCGCCATGTCGCCCCCGGCCG | 1 | AD | 1 | b38 | Y |
| diagnosed | SOX10 | 22:37983444:C:G | 1 | AD | 1 | b38 | Y |
| diagnosed | SOX10 | 22:37983391:C:G | 1 | AD | 2 | b38 | Y |
| diagnosed | SOX10 | 22:37983432:G:T | 1 | AD | 2 | b38 | N |
| diagnosed | SOX10 | 22:37978016:G:GA | 2 | AD | 1 | b38 | Y |
| diagnosed | SOX10 | 22:37983466:GC:G | 1 | AD | 1 | b38 | N |
| diagnosed | SOX10 | 22:37979330-37985133:DEL | 7 | AD | - | b38 | Y |
| partially ( HL explained ) | SOX10 | 22:37983390:G:A | 3 | AD | NA | b38 | N |
| diagnosed | SOX10 | 22:37983429:C:T | 1 | AD | 3 | b38 | Y |
| diagnosed | SOX11 | 2:5833008:C:T | 1 | AD | 1 | b37 | N |
| diagnosed | SOX11 | 2:5692878:A:G | 1 | AD | 1 | b38 | N |
| diagnosed | SPATA5 | 4:122934573:TCAA:T\|4:122947489:G:A | 1 | AR | 1 | b38 | N |
| diagnosed | SPATA5 | 4:122923143:A:C\|4:123256059:C:G | 2 | AR | 1 | b38 | N |
| diagnosed | SPATA5L1 | 15:45402956:G:T\|15:45418675:G:T | 13 | AR | 3 | b38 | N |
| diagnosed | SPATA5L1 | 15:45402956:G:T\|15:45418675:G:T | 4 | AR | 3 | b38 | N |
| diagnosed | SPATA5L1 | 15:45415617:GC:G\|15:45415624:T:C | 5 | AR | 3 | b38 | N |
| diagnosed | SPG7 | 16:89526449:C:T | 1 | AR | 1 | b38 | N |
| diagnosed | SPG7 | 16:89531962:G:GC\|16:89546737:C:T | 1 | AR | 1 | b38 | N |
| diagnosed | SPOP | 17:49619031:C:T | 3 | AD | 3 | b38 | N |
| diagnosed | SRCAP | 16:30737969:CCT:C | 1 | AD | 1 | b38 | N |
| diagnosed | ST3GAL5 | 2:85848164:C:A | 2 | AR | 2 | b38 | N |
| diagnosed | ST3GAL5 | 2:85848164:C:A | 1 | AR | 2 | b38 | N |
| diagnosed | STAC3 | 12:57244322:C:G | 1 | AR | 2 | b38 | N |
| diagnosed | STRC | 15:43601395:C:T | 1 | AR | 1 | b38 | Y |
| diagnosed | STRC | 15:43601395:C:T | 1 | AR | 1 | b38 | N |
| diagnosed | STRC | CNV | 1 | AR | - | | Y |
| diagnosed | STRC | 15:43597369-43605431:DEL | NA | AR | - | b38 | Y |
| diagnosed | STRC | 15:43560235-43695691 | NA | AR | - | b38 | Y |
| diagnosed | STRC | 15:43603276:TC:T\|CNV | 152 | AR | - | b38 | Y |
| diagnosed | STRC | 15:43600879:C:A\|15:43560148-43618624 | NA | AR | - | b38 | Y |
| diagnosed | STRC | 15:43560235-43695691:DEL\|15:43560148-43606091:DEL | NA | AR | - | b38 | Y |
| diagnosed | STRC | 15:43560148-43618624:DEL\|15:43560235-43695691:DEL | NA | AR | - | b38 | Y |
| diagnosed | STRC | 15:43560235-43695691 | NA | AR | - | b38 | Y |
| diagnosed | TBC1D24 | 16:2497067:A:C | 3 | AD | 2 | b38 | Y |
| diagnosed | TBC1D24 | 16:2496266:C:T\|16:2499421:G:A | 4 | AR | 1 | b38 | N |
| diagnosed | TBC1D32 | 6:121613198:C:T | 1 | AR | 3 | b37 | N |
| diagnosed | TBR1 | 2:161420224:C:T | 1 | AD | 1 | b38 | N |

| | | | | | | | |
|---|---|---|---|---|---|---|---|
| diagnosed | TCAP | 17:39665391:C:CG | 1 | AR | 1 | b38 | N |
| diagnosed | TECPR2 | 14:102909983:G:A\|14:102931627:G:C | 5 | AR | 3 | b37 | N |
| diagnosed | TECTA | 11:121168135:C:T | 1 | AD | 2 | b38 | Y |
| diagnosed | TECTA | 11:121038773:C:T | 1 | AD | 2 | b37 | Y |
| diagnosed | TECTA | 11:121028877:G:A\|11:121030843:G:A | 1 | AR | 1 | b37 | Y |
| diagnosed | TECTA | 11:121187831:G:T | 3 | AR | 1 | b38 | Y |
| diagnosed | TECTA | 11:121129989:C:T\|11:121168923:AG:A | 1 | AR | 1 | b38 | Y |
| partially ( HL explained ) | TECTA | 11:121137417:GCAGCA:G | 4 | AD | 1 | b38 | N |
| diagnosed | TECTA | 11:121168064:C:T | 1 | AD | 2 | b38 | Y |
| diagnosed | TECTA | 11:121187848:G:T | 1 | AD | NA | b38 | Y |
| diagnosed | TFAP2A | 6:10404634:G:T | 1 | AD | 3 | b38 | N |
| diagnosed | TFAP2A | 6:10404742:T:C | 1 | AD | NA | b37 | N |
| diagnosed | TFAP2A | 6:10404562:C:T | 1 | AD | 3 | b38 | N |
| diagnosed | TGDS | 13:94590868:C:A | 1 | AR | 2 | b38 | N |
| diagnosed | TGFBR1 | 9:99137980:G:C | 1 | AD | 2 | b38 | N |
| diagnosed | THOC6 | 16:3027379:G:A\|16:3027170:G:C\|16:3026140:T:A | 1 | AR | 2 | b38 | N |
| diagnosed | THOC6 | 16:3026140:T:A\|16:3027170:G:C\|16:3027379:G:A | 1 | AR | 2 | b38 | N |
| diagnosed | TMC1 | 9:72791995:G:A\|9:72820918:TG:T | 1 | AR | 1 | b38 | Y |
| diagnosed | TMC1 | 9:72694715:G:A\|9:72789207:G:A | 1 | AR | 1 | b38 | Y |
| diagnosed | TMC1 | 9:72789193:T:G\|9:72820841:G:C | 1 | AR | 1 | b38 | Y |
| diagnosed | TMC1 | 9:75431077:G:A | 1 | AD | 2 | b37 | Y |
| diagnosed | TMC1 | 9:72816161:G:A | 2 | AD | 3 | b38 | Y |
| diagnosed | TMEM260 | 14:57092111:C:G | 1 | AR | 3 | b37 | N |
| diagnosed | TMPRSS3 | 21:42389042:TG:T | 1 | AR | 1 | b38 | Y |
| diagnosed | TMPRSS3 | 21:42375844:A:G | 2 | AR | 2 | b38 | Y |
| diagnosed | TMPRSS3 | 21:42388402:C:A\|21:42388436:G:T | 1 | AR | 1 | b38 | Y |
| partially ( HL explained ) | TMPRSS3 | 21:42388532:C:T | 5 | AR | 2 | b38 | N |
| diagnosed | TMPRSS3 | 21:42389042:TG:T | 1 | AR | 1 | b38 | Y |
| diagnosed | TMPRSS3 | 21:42388436:G:T\|21:42389042:TG:T | 1 | AR | 1 | b38 | Y |
| diagnosed | TMPRSS3 | 21:42388524:G:A\|21:42389042:TG:T | 1 | AR | 1 | b38 | Y |
| diagnosed | TMPRSS3 | 21:42383991:C:T\|21:42388436:G:T | 2 | AR | 2 | b38 | Y |
| diagnosed | TMPRSS3 | 21:42382101:C:T\|21:42388935:G:A | 1 | AR | 2 | b38 | Y |
| diagnosed | TNFRSF11B | 8:118926720:T:C\|8:118951821:T:C | 1 | AR | 1 | b38 | N |
| diagnosed | TNPO2 | 19:12817396:T:A | 4 | AD | 3 | b37 | N |
| diagnosed | TP63 | 3:189864379:C:T | 1 | AD | 1 | b38 | N |
| diagnosed | TRNT1 | 3:3137406:C:T | 10 | AR | 2 | b38 | N |
| diagnosed | TRRAP | 7:98927284:T:G | 3 | AD | 3 | b38 | N |
| diagnosed | TUBB | 6:30722551:C:G | 2 | AD | 1 | b38 | N |
| diagnosed | TUBB4B | 9:140137742:C:T | 2 | AD | 3 | b37 | N |
| diagnosed | TXNL4A | 18:79988298:TGG:T | 20 | AD | 3 | b38 | N |
| diagnosed | TXNL4A | 18:79973859:G:C\|18:79988581-799886:DEL | NA | AR | - | b38 | N |
| diagnosed | UBE3A | CNV | NA | UPD | - | | N |
| diagnosed | UBE3B | 12:109948259:C:T | 1 | AR | 1 | b37 | N |
| diagnosed | USH1C | 11:17498251:C:A\|11:17531408:C:CG | 1 | AR | 1 | b38 | N |
| diagnosed | USH1C | 11:17531408:C:CG | 2 | AR | 1 | b38 | N |
| diagnosed | USH2A | 1:215848480:G:T\|1:216420436:TC:T | 1 | AR | 1 | b37 | Y |
| partially ( HL explained ) | USH2A | 1:215817143:C:A\|1:216199725:G:C | 1 | AR | 1 | b38 | Y |
| diagnosed | USH2A | 1:216048537:CT:C\|1:216247094:TC:T | 1 | AR | 1 | b38 | N |
| diagnosed | USH2A | 1:215891198:T:C\|1:216250931:G:A | 54 | AR | NA | b38 | Y |
| diagnosed | USH2A | 1:215877882:T:C\|1:215999016:A:T | 1 | AR | 1 | b38 | N |
| diagnosed | USH2A | 1:215848044:GCC:G\|1:215848514:C:T | 1 | AR | 1 | b37 | N |
| diagnosed | USH2A | 1:216046496:T:G\|1:216247094:TC:T | 1 | AR | 1 | b38 | N |
| diagnosed | USH2A | 1:215822026:G:A\|1:215931934:T:TA | 1 | AR | 2 | b37 | N |
| diagnosed | USH2A | 1:216247094:TC:T | 1 | AR | 1 | b38 | N |
| diagnosed | USH2A | 1:216420436:TC:T | 1 | AR | 1 | b37 | N |
| diagnosed | USH2A | 1:216250989:C:T\|1:216422150:G:A | 1 | AR | 1 | b38 | N |
| diagnosed | USH2A | 1:215648584:TGAGGCCAGCGTCCC:T\|1:216247094:TC:T | 1 | AR | 1 | b38 | N |
| diagnosed | USH2A | 1:215798991:GGCCATGGCCATCAT:G | 1 | AR | 1 | b38 | N |
| diagnosed | USH2A | 1:216247094:TC:T\|1:216325524:G:GTGGC | 1 | AR | 1 | b38 | N |
| diagnosed | USH2A | 1:215798991:GGCCATGGCCATCAT:G | 1 | AR | 1 | b38 | N |
| diagnosed | USH2A | 1:215759826:G:A\|1:216247094:TC:T | 1 | AR | 1 | b38 | N |
| diagnosed | USH2A | 1:215674536:AT:A | 1 | AR | 1 | b38 | N |
| diagnosed | USH2A | 1:215728383:G:A\|1:215758647:AT:A | 1 | AR | 1 | b38 | N |

| | | | | | | | |
|---|---|---|---|---|---|---|---|
| diagnosed | USH2A | 1:216247094:TC:T | 1 | AR | 1 | b38 | N |
| diagnosed | USH2A | 1:216247094:TC:T | 1 | AR | 1 | b38 | N |
| partially ( HL explained ) | USH2A | 1:215741377:G:C | 7 | AR | 1 | b38 | N |
| diagnosed | USH2A | 1:215741377:G:C | 1 | AR | 1 | b38 | N |
| diagnosed | USH2A | 1:216247094:TC:T\|1:216325524:G:GTGGC | 1 | AR | 1 | b38 | N |
| diagnosed | USH2A | 1:215891198:T:C | 1 | AR | NA | b38 | N |
| diagnosed | USH2A | 1:216070294:T:A | 1 | AR | 1 | b38 | N |
| diagnosed | USH2A | 1:215674536:AT:A\|1:215680252:G:T | 1 | AR | 1 | b38 | N |
| diagnosed | USH2A | 1:215639190:G:A\|1:216247094:TC:T | 1 | AR | 1 | b38 | N |
| diagnosed | USH2A | 1:215675619:A:T\|1:216247094:TC:T | 1 | AR | 1 | b38 | N |
| diagnosed | USH2A | 1:216247094:TC:T\|1:216325524:G:GTGGC | 1 | AR | 1 | b38 | N |
| diagnosed | USH2A | 1:216097126:AG:A\|1:216247094:TC:T | 1 | AR | 1 | b38 | N |
| diagnosed | USH2A | 1:216247094:TC:T | 1 | AR | 1 | b38 | N |
| diagnosed | USH2A | 1:215889057:G:C\|1:216321921:A:G | 1 | AR | 2 | b38 | N |
| diagnosed | USH2A | 1:216175368:C:CT\|1:216247094:TC:T | 1 | AR | 1 | b38 | Y |
| diagnosed | USH2A | 1:216073258:G:GATGCCAAGTTA\|1:216247118:C:A | 1 | AR | 3 | b38 | N |
| diagnosed | USH2A | 1:215817108:G:T\|1:215889057:G:C | 1 | AR | 1 | b38 | N |
| diagnosed | USH2A | 1:215674595:G:A\|1:216324240:C:A | 1 | AR | 2 | b38 | N |
| diagnosed | USH2A | 1:216078088:C:T\|1:216207280:G:T | 1 | AR | 1 | b38 | Y |
| diagnosed | USH2A | 1:216247094:TC:T\|1:216321897:T:G | 1 | AR | 1 | b38 | N |
| diagnosed | USH2A | 1:215674852:CTCCAGAGTTGTGA:C\|1:216000398:C:T | 1 | AR | 1 | b38 | Y |
| diagnosed | USH2A | 1:215901574:C:T\|1:216420436:TC:T | 1 | AR | 1 | b37 | Y |
| diagnosed | USH2A | 1:215675619:A:T\|1:215817108:G:T | 1 | AR | 1 | b38 | N |
| diagnosed | USH2A | 1:215798952:C:CT\|1:216247094:TC:T | 1 | AR | 1 | b38 | Y |
| partially ( HL explained ) | USH2A | 1:215675516:A:G\|1:215758613:CA:C | 1 | AR | 1 | b38 | Y |
| diagnosed | USH2A | 1:216420436:TC:T\|1:216595516:G:A | 1 | AR | 1 | b37 | Y |
| diagnosed | USH2A | 1:216380742:TTG:T\|1:216420436:TC:T | 1 | AR | 1 | b37 | N |
| diagnosed | USH2A | 1:216064540:T:C\|1:216420436:TC:T | 1 | AR | NA | b37 | Y |
| diagnosed | USH2A | 1:216172401:C:T\|1:216380742:TTG:T | 1 | AR | 1 | b37 | N |
| diagnosed | USH2A | 1:215848044:GCC:G\|1:216270538:G:A | 1 | AR | 1 | b37 | Y |
| diagnosed | USH2A | 1:215847598:C:T\|1:216062399:G:C | 1 | AR | 1 | b37 | Y |
| diagnosed | USH2A | 1:216420436:TC:T\|1:216497582:C:A | 1 | AR | 1 | b37 | Y |
| diagnosed | USH2A | 1:215848299:G:T\|1:216498866:G:GTGGC | 1 | AR | 1 | b37 | N |
| diagnosed | USH2A | 1:216270538:G:A | 1 | AR | 1 | b37 | Y |
| diagnosed | USH2A | 1:215848514:C:T\|1:215853549:TTC:T | 1 | AR | 1 | b37 | Y |
| diagnosed | USH2A | 1:216246438:C:T | 1 | AR | NA | b37 | N |
| diagnosed | USH2A | 1:215896963:G:C\|1:216498754:T:G | NA | AR | NA | b37 | Y |
| diagnosed | USH2A | 1:215782125:C:T\|1:215836457:215842264:DEL | 1 | AR | - | b38 | Y |
| diagnosed | USP9X | X:41152955:C:T | 1 | XD | 1 | b38 | N |
| diagnosed | VPS13B | 8:99642105:C:T | 1 | AR | 3 | b38 | N |
| diagnosed | WDR35 | 2:20133230:C:T\|2:20169255:G:A | 1 | AR | 1 | b37 | N |
| diagnosed | WDR45 | X:49075131:C:T | 3 | XD | NA | b38 | N |
| partially ( HL explained ) | WFS1 | 4:6301846:C:T | 1 | AD | 2 | b38 | Y |
| diagnosed | WFS1 | 4:6301035:TTCG:T\|4:6301680:C:T | 2 | AR | 2 | b38 | N |
| diagnosed | WFS1 | 4:6302303:G:C | 1 | AD | 1 | b38 | N |
| diagnosed | WFS1 | 4:6301846:C:T | 1 | AD | 2 | b38 | N |
| diagnosed | WFS1 | 4:6301846:C:T | 1 | AD | 1 | b38 | Y |
| diagnosed | WFS1 | 4:6301846:C:T | 1 | AD | 1 | b38 | Y |
| diagnosed | WFS1 | 4:6301903:G:A | 1 | AD | 2 | b38 | Y |
| diagnosed | WFS1 | 4:6301846:C:T | 1 | AD | 1 | b38 | Y |
| diagnosed | WFS1 | 4:6291229:C:T\|4:6302001:G:C | 1 | AR | 1 | b38 | N |
| diagnosed | WFS1 | 4:6289068:G:A\|4:6302449:C:T | 3 | AR | 2 | b38 | N |
| diagnosed | WFS1 | 4:6301846:C:T | 1 | AD | 2 | b38 | N |
| diagnosed | WFS1 | 4:6303573:C:T | 1 | AD | 2 | b37 | Y |
| diagnosed | ZBTB20 | 3:114351344:G:C | 2 | AD | 3 | b38 | Y |
| diagnosed | ZBTB20 | 3:114058130:T:G | 1 | AD | NA | b37 | N |
| diagnosed | ZEB2 | 2:144389918:A:G | 2 | AD | 2 | b38 | N |
| diagnosed | ZMIZ1 | 10:81056377:G:GC | 1 | AD | 3 | b37 | N |
| diagnosed | ZMIZ1 | 10:79311069:CA:C | 1 | AD | 3 | b38 | N |

**Supplementary Table 2: Additional Information on diagnoses observed within HL and HED Cohorts**
This table provides supplementary details on the 625 diagnoses identified in the broader cohort defined by HPO terms 211 of which belong to the "Hearing and Ear Disorders" (HED) cohort. The columns include information on whether the case is solved the diagnosed gene and associated variants Exomiser rank and mode of inheritance (MOI) tiering genome assembly version and a flag indicating inclusion in the HED cohort.
