## Supplementary Table 3 for "The genomic landscape of syndromic and non-syndromic hearing loss within the 100,000 Genomes Project cohort"

| Stratification variable | Variable | HL cohort | | HED | |
|---|---|---|---|---|---|
| | | freq / tot freq | Solve rate (%) | freq / tot freq | Solve rate (%) |
| onset | Adulthood | 9 / 54 | 16.66666667 | 4 / 26 | 15.38461538 |
| | Childhood | 66 / 253 | 26.08695652 | 36 / 138 | 26.08695652 |
| | Congenital | 123 / 370 | 33.24324324 | 97 / 300 | 32.33333333 |
| severity | Mild | 40 / 150 | 26.66666667 | 9 / 44 | 20.45454545 |
| | Moderate | 58 / 216 | 26.85185185 | 33 / 125 | 26.4 |
| | Profound | 54 / 176 | 30.68181818 | 42 / 138 | 30.43478261 |
| | Severe | 75 / 256 | 29.296875 | 56 / 185 | 30.27027027 |
| laterality | Bilateral | 199 / 738 | 26.96476965 | 144 / 526 | 27.37642586 |
| | Unilateral | 14 / 126 | 11.11111111 | 2 / 23 | 8.695652174 |
| frequency | High frequency | 78 / 241 | 32.36514523 | 68 / 198 | 34.34343434 |
| | Low frequency | 5 / 38 | 13.15789474 | 2 / 23 | 8.695652174 |
| | Mid frequency | 8 / 47 | 17.0212766 | 5 / 31 | 16.12903226 |
| family structure | 1 | 137 / 565 | 24.24778761 | 31 / 133 | 23.30827068 |
| | 2 | 121 / 448 | 27.00892857 | 38 / 137 | 27.73722628 |
| | 3 | 311 / 1090 | 28.53211009 | 106 / 408 | 25.98039216 |
| | 4 | 48 / 146 | 32.87671233 | 31 / 81 | 38.27160494 |
| | 5+ | 8 / 22 | 36.36363636 | 5 / 15 | 33.33333333 |
| syndromic | non-syndromic | 130 / 500 | 26 | 129 / 492 | 26.2195122 |
| | syndromic | 495 / 1771 | 27.95031056 | 82 / 282 | 29.07801418 |
| type | Conductive | 61 / 254 | 24.01574803 | 5 / 45 | 11.11111111 |
| | Mixed | 4 / 29 | 13.79310345 | 1 / <5 | ≥ 25 |
| | Sensorineural | 394 / 1312 | 30.0304878 | 196 / 683 | 28.69692533 |
| progression | Non-Progressive | 7 / 24 | 29.16666667 | 5 / 13 | 38.46153846 |
| | Progressive | 51 / 209 | 24.40191388 | 44 / 174 | 25.28735632 |

**Supplementary Table 3: Stratified solve rates for HL and HED cohorts**
This table presents the solve rates for different stratification variables within the HL and HED cohorts, including onset, severity, laterality, frequency, family structure, syndromic status, hearing loss type and progression. Solve rates are expressed as relative frequencies and corresponding percentages, representing the proportion of successfully diagnosed cases within each category. To ensure compliance with data protection policies, any frequency counts lower than 5 associated with phenotypic information have been redacted and are represented as '<5' and the minimum possible solve rate corresponding to the lowest redacted count is shown.
