## Supplementary Table 4 for "The genomic landscape of syndromic and non-syndromic hearing loss within the 100,000 Genomes Project cohort"

| Stratification variable | Variable tested combination | HL cohort | | | | HED cohort | | | |
|---|---|---|---|---|---|---|---|---|---|
| | | or | pvalue | CI | significance pvalue | or | pvalue | CI | significance pvalue |
| onset | Adulthood;Childhood | 1.764705882 | 0.165164663 | 0.794215806042931-4.32707024560104 | | 1.941176471 | 0.322823058 | 0.598688321120605-8.24358211795237 | |
| | Adulthood;Congenital | 2.489878543 | 0.017456537 | 1.15159331121699-5.97534376315967 | * | 2.628078818 | 0.080130985 | 0.857295668644363-10.7544056980767 | |
| | Childhood;Congenital | 1.410931174 | 0.062491519 | 0.976400648381555-2.04736874638044 | | 1.353858785 | 0.21864672 | 0.845991084080964-2.1926480438373 | |
| severity | Mild;Moderate | 1.009493671 | 1 | 0.615002738797159-1.66683436771926 | | 1.394927536 | 0.544035301 | 0.576142091870476-3.65355448258952 | |
| | Mild;Profound | 1.217213115 | 0.462799084 | 0.730558384879592-2.03637305648795 | | 1.701388889 | 0.248842595 | 0.71755722659204-4.37985812740904 | |
| | Mild;Severe | 1.139502762 | 0.648232027 | 0.71034406455152-1.84295305350793 | | 1.688199828 | 0.264048886 | 0.731525796366098-4.2575185138675 | |
| | Moderate;Profound | 0.829348336 | 0.432347519 | 0.521973719159595-1.31959310296135 | | 1.21969697 | 0.496419133 | 0.688214359656131-2.17025735048426 | |
| | Moderate;Severe | 1.128786436 | 0.607919266 | 0.739461572107897-1.72743637291234 | | 1.210241954 | 0.522721602 | 0.709233407450173-2.08300757424449 | |
| | Profound;Severe | 0.936157152 | 0.830635882 | 0.603744768196038-1.45678861939392 | | 0.992248062 | 1 | 0.598135045631252-1.65266198766358 | |
| laterality | Bilateral;Unilateral | 2.953617811 | 7.61E-05 | 1.63859157917452-5.70437083091953 | *** | 3.958115183 | 0.053114607 | 0.946053864363848-35.1890953284557 | |
| frequency | High frequency;Low frequency | 3.158282209 | 0.020667881 | 1.15858659185231-10.7297221884239 | * | 5.492307692 | 0.015845709 | 1.27265216276392-49.4359417228996 | * |
| | High frequency;Mid frequency | 2.332822086 | 0.037253934 | 1.00935131782372-6.04403563323904 | * | 2.72 | 0.060458109 | 0.964666660085538-9.44788853467777 | |
| | Low frequency;Mid frequency | 1.353846154 | 0.764992636 | 0.349078988233634-5.77299194643536 | | 2.019230769 | 0.685350995 | 0.289604341777488-22.9536641353743 | |
| family structure | 1;2 | 0.865045957 | 0.345311579 | 0.645008337158798-1.16121635291652 | | 1.262952102 | 0.485525577 | 0.703665873760705-2.2754720281195 | |
| | 1;3 | 1.247228807 | 0.070455329 | 0.982466110698501-1.58762837733331 | | 1.154881436 | 0.567891639 | 0.71690329760266-1.89445672470382 | |
| | 1;4 | 0.653524143 | 0.043974973 | 0.433487935138535-0.994493799489773 | * | 0.490196078 | 0.029111824 | 0.25721724221074-0.937249438868974 | * |
| | 1;5+ | 0.560163551 | 0.209963214 | 0.214147761819471-1.57748912849111 | | 0.607843137 | 0.362034686 | 0.173665881506912-2.45095256244307 | |
| | 2;3 | 1.078910237 | 0.574372497 | 0.838012758790692-1.39366345642004 | | 0.914430115 | 0.737035648 | 0.581679935994388-1.45564063063552 | |
| | 2;4 | 0.755479103 | 0.171865915 | 0.496452485785082-1.15934181624732 | | 0.619094167 | 0.131772003 | 0.332059605653127-1.16044927616745 | |
| | 2;5+ | 0.647553517 | 0.334916057 | 0.246465742214059-1.83037808673695 | | 0.767676768 | 0.763285498 | 0.221628208751311-3.06010263568963 | |
| | 3;4 | 0.815094138 | 0.286285737 | 0.556357520904187-1.20683063118033 | | 0.566118351 | 0.03000125 | 0.334840397415466-0.969594310495851 | * |
| | 3;5+ | 0.698652118 | 0.475436014 | 0.270464918035424-1.94295982142166 | | 0.701986755 | 0.553055438 | 0.212919249431296-2.68138322673658 | |
| | 4;5+ | 0.857142857 | 0.80967009 | 0.310339438369457-2.53121990245317 | | 1.24 | 0.779884286 | 0.345192895443965-5.06132083780363 | |
| syndromic | non-syndromic;syndromic | 1.104111406 | 0.395801711 | 0.877410358857074-1.39472321429238 | | 0.866760734 | 0.40239978 | 0.61808429325112-1.21939941183039 | |
| type | Conductive;Mixed | 1.975388601 | 0.252669905 | 0.644345324550415-8.09929053084005 | | 0.375 | 0.417536672 | 0.0244999690870151-23.5820281381469 | |
| | Conductive;Sensorineural | 1.357941355 | 0.058959249 | 0.987347261434358-1.88638535803528 | | 3.219712526 | 0.00928811 | 1.24328849722317-10.5945888549299 | ** |
| | Mixed;Sensorineural | 2.682461874 | 0.06475636 | 0.917153165000758-10.6715451059781 | | 1.207392197 | 1 | 0.0962517284907923-63.685747368446 | |
| progression | Non-Progressive;Progressive | 0.783905967 | 0.62145248 | 0.28882743820431-2.36902978272953 | | 0.541538462 | 0.330507757 | 0.147616363184355-2.22643032141352 | |

**Supplementary Table 4: Two-sided Fisher’s exact test comparative analysis of solve rates in stratified cohorts**
This table presents the results of a comparative analysis of solve rates across various stratification variables within the HL and HED cohorts. The table includes odds ratios (OR), p-values, and confidence intervals (CI) for different variable combinations related to onset, severity, laterality, frequency, family structure, syndromic status, hearing loss type, and progression. The * indicates p values < 0.05 after two-sided Fisher’s exact test, ** p values <0.01 and *** p values <0.001 as in FIgure 3.
