## Supplementary Table 5 for "The genomic landscape of syndromic and non-syndromic hearing loss within the 100,000 Genomes Project cohort"

| Phenotypic abnormalities combination | Diagnosed gene | Freq |
|---|---|---|
| Abnormality of the ear | GJB2 | 22 |
| Abnormality of the ear | MYO15A | 10 |
| Abnormality of the ear | USH2A | 8 |
| Abnormality of the ear | STRC | 7 |
| Abnormality of the ear | TMPRSS3 | 7 |
| Abnormality of the ear | SLC26A4 | 6 |
| Abnormality of the ear | TECTA | 6 |
| Abnormality of the ear | MYO6 | 5 |
| Abnormality of the ear | TMC1 | 5 |
| Abnormality of the ear | CDH23 | <5 |
| Abnormality of the ear | MYO7A | <5 |
| Abnormality of the ear | SOX10 | <5 |
| Abnormality of the ear | GATA3 | <5 |
| Abnormality of the ear | LOXHD1 | <5 |
| Abnormality of the ear | MITF | <5 |
| Abnormality of the ear | OTOF | <5 |
| Abnormality of the ear | EYA4 | <5 |
| Abnormality of the ear | GSDME | <5 |
| Abnormality of the ear | OTOG | <5 |
| Abnormality of the ear | ADGRV1 | <5 |
| Abnormality of the ear | ATP6V1B2 | <5 |
| Abnormality of the ear | BSND | <5 |
| Abnormality of the ear | CLIC5 | <5 |
| Abnormality of the ear | CLRN1 | <5 |
| Abnormality of the ear | COCH | <5 |
| Abnormality of the ear | COL11A2 | <5 |
| Abnormality of the ear | EPS8L2 | <5 |
| Abnormality of the ear | ESPN | <5 |
| Abnormality of the ear | FBP1 | <5 |
| Abnormality of the ear | GRXCR1 | <5 |
| Abnormality of the ear | KCNQ4 | <5 |
| Abnormality of the ear | LHFPL5 | <5 |
| Abnormality of the ear | MARVELD2 | <5 |
| Abnormality of the ear | OTOA | <5 |
| Abnormality of the ear | PAX3 | <5 |
| Abnormality of the ear | PDZD7 | <5 |
| Abnormality of the ear | PJVK | <5 |
| Abnormality of the ear | PLS1 | <5 |
| Abnormality of the ear | PTPN11 | <5 |
| Abnormality of the ear | SIX1 | <5 |
| Abnormality of the ear | SMPX | <5 |
| Abnormality of the ear | TBC1D24 | <5 |
| Abnormality of the ear | WFS1 | <5 |
| Abnormality of the ear, Abnormality of the eye | USH2A | 37 |
| Abnormality of the ear, Abnormality of the eye | MYO7A | 8 |
| Abnormality of the ear, Abnormality of the eye | WFS1 | 6 |
| Abnormality of the ear, Abnormality of the eye | CDH23 | <5 |
| Abnormality of the ear, Abnormality of the eye | CLRN1 | <5 |
| Abnormality of the ear, Abnormality of the eye | PRPS1 | <5 |
| Abnormality of the ear, Abnormality of the eye | USH1C | <5 |
| Abnormality of the ear, Abnormality of the eye | ABHD12 | <5 |
| Abnormality of the ear, Abnormality of the eye | ADGRV1 | <5 |
| Abnormality of the ear, Abnormality of the eye | CACNA1G | <5 |
| Abnormality of the ear, Abnormality of the eye | COL11A1 | <5 |
| Abnormality of the ear, Abnormality of the eye | CYP1B1 | <5 |
| Abnormality of the ear, Abnormality of the eye | GJB2 | <5 |

| | | |
|---|---|---|
| Abnormality of the ear, Abnormality of the eye | GUCY2D | <5 |
| Abnormality of the ear, Abnormality of the eye | LRP2 | <5 |
| Abnormality of the ear, Abnormality of the eye | MIP\|WFS1 | <5 |
| Abnormality of the ear, Abnormality of the eye | MITF | <5 |
| Abnormality of the ear, Abnormality of the eye | MYO15A | <5 |
| Abnormality of the ear, Abnormality of the eye | PHYH | <5 |
| Abnormality of the ear, Abnormality of the eye | STRC | <5 |
| Abnormality of the ear, Abnormality of the eye | TECTA | <5 |
| Abnormality of the ear, Abnormality of the eye | TFAP2A | <5 |
| Abnormality of the ear, Abnormality of the nervous system | GJB2 | 5 |
| Abnormality of the ear, Abnormality of the nervous system | MYO6 | <5 |
| Abnormality of the ear, Abnormality of the nervous system | ATP1A3 | <5 |
| Abnormality of the ear, Abnormality of the nervous system | EIF2B5 | <5 |
| Abnormality of the ear, Abnormality of the nervous system | EIF3F | <5 |
| Abnormality of the ear, Abnormality of the nervous system | EYA4 | <5 |
| Abnormality of the ear, Abnormality of the nervous system | FGF3 | <5 |
| Abnormality of the ear, Abnormality of the nervous system | FOXC1 | <5 |
| Abnormality of the ear, Abnormality of the nervous system | KAT6A | <5 |
| Abnormality of the ear, Abnormality of the nervous system | KAT6B | <5 |
| Abnormality of the ear, Abnormality of the nervous system | KCNQ4 | <5 |
| Abnormality of the ear, Abnormality of the nervous system | MN1 | <5 |
| Abnormality of the ear, Abnormality of the nervous system | NF2 | <5 |
| Abnormality of the ear, Abnormality of the nervous system | PPP1R12A | <5 |
| Abnormality of the ear, Abnormality of the nervous system | SETD5 | <5 |
| Abnormality of the ear, Abnormality of the nervous system | SOX10 | <5 |
| Abnormality of the ear, Abnormality of the nervous system | SRCAP | <5 |
| Abnormality of the ear, Abnormality of the nervous system | THOC6 | <5 |
| Abnormality of the ear, Abnormality of the nervous system | USH2A | <5 |
| Abnormality of the ear, Abnormality of the nervous system | WDR45 | <5 |
| Abnormality of the ear, Abnormality of the eye, Abnormality of the nervous system | OPA1 | <5 |
| Abnormality of the ear, Abnormality of the eye, Abnormality of the nervous system | AIFM1 | <5 |
| Abnormality of the ear, Abnormality of the eye, Abnormality of the nervous system | ATP1A3 | <5 |
| Abnormality of the ear, Abnormality of the eye, Abnormality of the nervous system | BBS1 | <5 |
| Abnormality of the ear, Abnormality of the eye, Abnormality of the nervous system | DNMT1\|PRKCG | <5 |
| Abnormality of the ear, Abnormality of the eye, Abnormality of the nervous system | HSD17B4 | <5 |
| Abnormality of the ear, Abnormality of the eye, Abnormality of the nervous system | LRP2 | <5 |
| Abnormality of the ear, Abnormality of the eye, Abnormality of the nervous system | PEX11B | <5 |
| Abnormality of the ear, Abnormality of the eye, Abnormality of the nervous system | PITX3 | <5 |
| Abnormality of the ear, Abnormality of the eye, Abnormality of the nervous system | POLG | <5 |
| Abnormality of the ear, Abnormality of the eye, Abnormality of the nervous system | PRPS1 | <5 |
| Abnormality of the ear, Abnormality of the eye, Abnormality of the nervous system | SALL1 | <5 |
| Abnormality of the ear, Abnormality of the eye, Abnormality of the nervous system | SLC52A2 | <5 |
| Abnormality of the ear, Abnormality of the eye, Abnormality of the nervous system | SPATA5L1 | <5 |
| Abnormality of the ear, Abnormality of the genitourinary system | COL4A3 | <5 |
| Abnormality of the ear, Abnormality of the genitourinary system | COL4A5 | <5 |
| Abnormality of the ear, Abnormality of the genitourinary system | PKD2 | <5 |
| Abnormality of the ear, Abnormality of the genitourinary system | SALL1 | <5 |
| Abnormality of the ear, Abnormality of the genitourinary system | TMPRSS3 | <5 |
| Abnormality of the ear, Abnormality of the musculature, Abnormality of the nervous system | AARS1 | <5 |
| Abnormality of the ear, Abnormality of the musculature, Abnormality of the nervous system | CAMTA1 | <5 |
| Abnormality of the ear, Abnormality of the musculature, Abnormality of the nervous system | CHAMP1 | <5 |
| Abnormality of the ear, Abnormality of the musculature, Abnormality of the nervous system | GJB2 | <5 |
| Abnormality of the ear, Abnormality of the musculature, Abnormality of the nervous system | MPV17 | <5 |
| Abnormality of the ear, Abnormality of the musculature, Abnormality of the nervous system | UBE3A | <5 |
| Abnormality of the ear, Abnormality of the nervous system, Growth abnormality | HDAC8 | <5 |
| Abnormality of the ear, Abnormality of the nervous system, Growth abnormality | AHDC1 | <5 |
| Abnormality of the ear, Abnormality of the nervous system, Growth abnormality | CDK5RAP2 | <5 |

| | | |
|---|---|---|
| Abnormality of the ear, Abnormality of the nervous system, Growth abnormality | LMNB2 | <5 |
| Abnormality of the ear, Abnormality of the nervous system, Growth abnormality | MORC2 | <5 |
| Abnormality of the ear, Abnormality of the nervous system, Growth abnormality | PTPN11 | <5 |
| Abnormality of the ear, Abnormality of the nervous system, Growth abnormality | PUS7 | <5 |
| Abnormality of head or neck, Abnormality of the ear | COL11A1 | <5 |
| Abnormality of head or neck, Abnormality of the ear | OTX2 | <5 |
| Abnormality of head or neck, Abnormality of the ear | POU3F4 | <5 |
| Abnormality of head or neck, Abnormality of the ear | TECTA | <5 |
| Abnormality of the cardiovascular system, Abnormality of the ear | EYA1 | <5 |
| Abnormality of the cardiovascular system, Abnormality of the ear | GLA | <5 |
| Abnormality of the cardiovascular system, Abnormality of the ear | KCNE1 | <5 |
| Abnormality of the cardiovascular system, Abnormality of the ear | KCNQ1 | <5 |
| Abnormality of the cardiovascular system, Abnormality of the ear | LOXHD1 | <5 |
| Abnormality of the cardiovascular system, Abnormality of the ear | MYBPC3\|TMPRSS3 | <5 |
| Abnormality of the ear, Abnormality of the nervous system, Abnormality of the skeletal system | ACTB | <5 |
| Abnormality of the ear, Abnormality of the nervous system, Abnormality of the skeletal system | EYA1 | <5 |
| Abnormality of the ear, Abnormality of the nervous system, Abnormality of the skeletal system | GNB1 | <5 |
| Abnormality of the ear, Abnormality of the nervous system, Abnormality of the skeletal system | KCNJ10 | <5 |
| Abnormality of the ear, Abnormality of the nervous system, Abnormality of the skeletal system | MYO15A | <5 |
| Abnormality of the ear, Abnormality of the nervous system, Abnormality of the skeletal system | SHANK3 | <5 |
| Abnormality of limbs, Abnormality of the ear, Abnormality of the musculature, Abnormality of the nervous system | EGR2 | <5 |
| Abnormality of limbs, Abnormality of the ear, Abnormality of the musculature, Abnormality of the nervous system | MORC2 | <5 |
| Abnormality of metabolism/homeostasis, Abnormality of the ear, Abnormality of the genitourinary system | COL4A5 | <5 |
| Abnormality of metabolism/homeostasis, Abnormality of the ear, Abnormality of the genitourinary system | MYO6 | <5 |
| Abnormality of head or neck, Abnormality of the ear, Abnormality of the nervous system, Abnormality of the skeletal system | ASXL2 | <5 |
| Abnormality of head or neck, Abnormality of the ear, Abnormality of the nervous system, Abnormality of the skeletal system | KIF1A | <5 |
| Abnormality of head or neck, Abnormality of the ear, Abnormality of the nervous system, Abnormality of the skeletal system | OTOG | <5 |
| Abnormality of the ear, Abnormality of the eye, Abnormality of the musculature, Abnormality of the nervous system | MME | <5 |
| Abnormality of the ear, Abnormality of the eye, Abnormality of the musculature, Abnormality of the nervous system | MYO7A | <5 |
| Abnormality of the ear, Abnormality of the eye, Abnormality of the musculature, Abnormality of the nervous system | PNPT1 | <5 |
| Abnormality of the ear, Abnormality of the eye, Abnormality of the musculature, Abnormality of the nervous system | SPATA5L1 | <5 |
| Abnormality of the ear, Abnormality of the eye, Abnormality of the musculature, Abnormality of the nervous system | USH2A | <5 |
| Abnormality of the ear, Abnormality of the skeletal system | COL1A2 | <5 |
| Abnormality of the ear, Abnormality of the skeletal system | COL1A1 | <5 |
| Abnormality of the ear, Abnormality of the skeletal system | TNFRSF11B | <5 |
| Abnormality of the ear, Abnormality of the skeletal system | USH2A | <5 |
| Abnormality of head or neck, Abnormality of the ear, Abnormality of the eye, Abnormality of the nervous system | ADNP | <5 |
| Abnormality of head or neck, Abnormality of the ear, Abnormality of the eye, Abnormality of the nervous system | CHD7 | <5 |
| Abnormality of head or neck, Abnormality of the ear, Abnormality of the eye, Abnormality of the nervous system | FGFR3 | <5 |
| Abnormality of head or neck, Abnormality of the ear, Abnormality of the nervous system | ADNP | <5 |
| Abnormality of head or neck, Abnormality of the ear, Abnormality of the nervous system | ANKRD11 | <5 |
| Abnormality of head or neck, Abnormality of the ear, Abnormality of the nervous system | COL11A2 | <5 |
| Abnormality of head or neck, Abnormality of the ear, Abnormality of the nervous system | SOX11 | <5 |
| Abnormality of head or neck, Abnormality of the ear, Abnormality of the nervous system | ZEB2 | <5 |
| Abnormality of the ear, Abnormality of the eye, Abnormality of the nervous system, Abnormality of the skeletal system | ANKRD11 | <5 |
| Abnormality of the ear, Abnormality of the eye, Abnormality of the nervous system, Abnormality of the skeletal system | ATP1A3 | <5 |
| Abnormality of the ear, Abnormality of the eye, Abnormality of the nervous system, Abnormality of the skeletal system | HIKESHI | <5 |
| Abnormality of the ear, Abnormality of the eye, Abnormality of the nervous system, Abnormality of the skeletal system | TBC1D24 | <5 |
| Abnormality of the cardiovascular system, Abnormality of the ear, Abnormality of the genitourinary system | COL4A4 | <5 |
| Abnormality of the cardiovascular system, Abnormality of the ear, Abnormality of the genitourinary system | COL4A5 | <5 |
| Abnormality of the cardiovascular system, Abnormality of the ear, Abnormality of the genitourinary system | PKD2 | <5 |
| Abnormality of the cardiovascular system, Abnormality of the ear, Abnormality of the genitourinary system | TMEM260 | <5 |

**Supplementary Figure 5: Diagnosed genes per phenotypic abnormality combination**
This table provides an overview of the diagnosed genes associated with various phenotypic abnormality combinations shown in Figure 4, including the frequency of each gene within the respective combinations. To ensure compliance with data protection policies, any frequency counts lower than 5 associated with phenotypic information have been redacted and are represented as '<5'.
